# The impact of contact tracing and household bubbles on deconfinement strategies for COVID-19: an individual-based modelling study

**DOI:** 10.1101/2020.07.01.20144444

**Authors:** Lander Willem, Steven Abrams, Pieter J. K. Libin, Pietro Coletti, Elise Kuylen, Oana Petrof, Signe Møgelmose, James Wambua, Sereina A. Herzog, Christel Faes, Philippe Beutels, Niel Hens

**Affiliations:** Centre for Health Economic Research and Modelling Infectious Diseases, Vaccine & Infectious Disease Institute, University of Antwerp, Antwerp, Belgium; Data Science Institute, Interuniversity Institute of Biostatistics and statistical Bioinformatics, UHasselt, Hasselt, Belgium; Global Health Institute, Department of Epidemiology and Social Medicine, University of Antwerp, Antwerp, Belgium; Centre for Population, Family and Health, University of Antwerp, Antwerp, Belgium; School of Public Health and Community Medicine, The University of New South Wales, Sydney, Australia

**Keywords:** infectious diseases, transmission dynamics, individual-based model, agent-based model, social distancing, behavioral changes, SARS-CoV-2, social contact patterns

## Abstract

**Background:** The COVID-19 pandemic caused many governments to impose policies restricting social interactions. These policies have slowed down the spread of the SARS-CoV-2 virus to the extent that restrictions were gradually lifted. Critical assessment of potential deconfinement strategies with respect to business, educational and leisure activities requires extensive scenario analyses.

**Methods:** We adapted the individual-based model “STRIDE” to simulate interactions between the 11 million inhabitants of Belgium at the levels of households, workplaces, schools and communities. We calibrated our model to the observed hospital incidence data, initial doubling time and serial seroprevalence data. STRIDE enables contact tracing and repetitive leisure contacts in extended household settings (so called “household bubbles”) with varying levels of connectivity.

**Results:** Household bubbles have the potential to reduce the number of COVID-19 hospital admissions by up to 90%. The effectiveness of contact tracing depends on its timing and the 4 days after the index case developed symptoms are crucial. The susceptibility of children affects the impact of a (partial) reopening of schools, though we found that social mixing patterns related to business and leisure activities are driving the COVID-19 burden.

**Conclusions:** Next to the absolute number and intensity of physical contacts, also their repetitiveness impacts the transmission dynamics and COVID-19 burden. The combination of closed networks and contact tracing seems essential for a controlled and persistent release of lockdown measures, but requires timely compliance to the bubble concept, testing, reporting and self-isolation.

## Introduction

As the COVID-19 pandemic rose, there was an urgent need to understand the transmission dynamics and potential impact of COVID-19 on healthcare capacity and to translate these insights into policy. Mathematical modelling has been essential to inform decision-making by estimating the consequences of unmitigated spread in the initial phase as well as the impact of non-pharmaceutical interventions. Gradually releasing society’s lockdown while keeping the spread of the virus under control, requires detailed models to simulate the (non-)propagation of SARS-Cov-2. To this end, it is important to capture the heterogeneity in social encounters by accounting for a low number of intense contacts (e.g., between household members) and a high(er) number of more fleeting contacts (e.g., during leisure activities, commuting, or in shops) [1].

Transmission models at the level of the individual allow for flexibility to cope with chance, age and context, which is especially of interest to study exit strategies involving school, workplace, leisure activities and microscale policies [2, 3]. Individual-based models (IBMs) pose a high burden on data-requirements, implementation and computation, however, the increasing availability of individual-level data facilitates thorough evaluation of specific intervention measures.

Understanding the interplay between human behavior and infectious disease dynamics is key to improve modelling and control efforts [4]. Social contact data has become available for numerous countries [5, 6] and has proven to be an invaluable source of information on the transmission of close contact infectious diseases [7, 8]. Social contact patterns can be used as a proxy for transmission dynamics when relying on the “social contact hypothesis” [7]. Disease-related proportionality factors and timings enable matching age-specific mixing patterns with observed incidence, prevalence, generation interval and reproduction number. Social contact patterns in a transmission model can be adjusted to simulate behavioural change and assess possible intervention strategies [4].

Given the rising number of confirmed COVID-19 cases and hospital admissions in Belgium during the beginning of March 2020, all schools, universities, cultural activities, bars and restaurants were closed from March 14th onward. Additional measures were imposed on March18th, with only work-related transport of essential workers allowed, and teleworking made the norm (termed “lockdown light” in comparison with more strict lockdowns in other countries). Hospital admissions peaked at the beginning of April, and declined afterwards [9]. Restrictive measures were gradually lifted from May 4th onward in terms of business-to-business (B2B), school, business-to-costumers (B2C) and leisure activities. There remains substantial uncertainty on the extent to which people complied with physical distancing guidelines during the deconfinement and how public awareness and interventions modified social contact characteristics. More specifically, did people mix in specific clusters and what was the effect of keeping distance, increased hygiene measures and wearing face masks? The nature of social contacts before and after the lockdown undoubtedly changed, and this affects the proportionality factors linking “contacts” with “transmission”. Prior to the SARS-CoV-2 pandemic, simulation models for infectious diseases could rely on documented social contact behavior as key input to model transmission dynamics. For COVID-19 predictions, there is however structural uncertainty on future social contact behavior, implying that additional runs or improved parameter estimation would not reduce it. For example, the incremental effect when contact tracing is in place depends on the tendency of people to meet others. If the population stays put, the effect of contact tracing is minimal because it would be dominated by the effect of having only within-household mixing, and the epidemic would fade out. This structural uncertainty can be captured through different social mixing assumptions within each strategy assessment.

In what follows, we analyse the effect of repetitive leisure contacts in extended household settings (so called “household bubbles”) on the transmission of COVID-19 and explore contact tracing strategies with respect to coverage, sensitivity and timing. Our analyses are based on the open-source IBM “STRIDE”, fitted to COVID-19 data from Belgium, with particular focus on transmission dynamics from adaptive social contact patterns.

## Methods

### Model structure

This work builds on a stochastic individual-based model (IBM) we developed for influenza [10, 11] and measles [12]. Our model is representative for the population of Belgium, covering 11 million unique individuals, runs in discrete time steps of 1 day while accounting for adjusted social contact patterns during weekdays, weekends, holiday periods, illness and the influence of public awareness and imposed policy measures. More details on the model structure, population, social contact patterns and stochastic realisations are provided in the Supplementary Material.

### Disease natural history

The health states in the IBM follow the conventional stages of susceptible, exposed, infectious and recovered, with the infectious health state divided in pre-symptomatic, symptomatic and asymptomatic. For every infected individual, we sample the onset and duration of each stage based on the distributions in Table S2.

### Social contact patterns

Social contact patterns for healthy, pre- and asymptomatic individuals are parameterized by a diary-based study performed in Belgium in 2010-2011 [13, 14, 15]. Contact rates at school and at work are conditional on school enrolment and employment, respectively. We account for behavioral changes of symptomatic cases using observations made during the 2009 H1N1 influenza pandemic in the UK [16], by reducing presence at school and work with 90%. Based on the same study, we reduce community engagement with 75% when experiencing symptoms. Transmission-relevant contact behavior within the household is assumed not to change when a household member develops symptoms.

### Parameter estimation

We estimated transmission and lockdown characteristics based on reported hospital admissions [9], initial doubling time (i.e., before the lockdown) [17] and serial seroprevalence data [18] up to May 1st. Afterwards, multiple restrictive measures in Belgium were relaxed, which is the focus of our scenario analysis. Details on the model parameters and our multi-criteria iterative procedure are provided in the Supplementary Material. Our iterative estimation procedure resulted in an ensemble of parameter sets that match our three reference criteria. From this ensemble, we selected a single best parameter set based on the average log-likelihood function value to match the observed hospital admissions over time, since this is the model outcome of main interest. The per-case average number of secondary cases in a susceptible population, which corresponds to the basic reproduction number R_0_, was estimated to be 3.42, which is in line with estimates from a meta-analysis [19] and other modelling studies for Belgium [20, 21]. Within our final model parameter ensemble, the reproduction number ranged between (3.41–3.49). The transmission model starts with 263 (236–307) infected cases on February 17th. The hospital probability for symptomatic cases over 80 years is 40% (35%–46%). From March 14th onward, the social contacts related to B2B decreased linearly to 14% (7%–30%) over 7 (5– 7) days. Contacts in the community during lockdown decreased to 15% (13%–18%) of pre-lockdown contact levels after 7 (5–7) days.

### Household bubbles

We defined a “household bubble” as a unique combination of 2 households in which the oldest household members cannot differ more than 3 years in age and are linked via their community contacts during weekends. The age-specific component is included to reduce inter-generational mixing, which is subject of sensitivity analyses with age-differences of 20 and 60 years. The assignment of household bubbles in STRIDE proceeds in a random order and if nomatching household is available, the household is not assigned to any household bubble. This procedure enables us to assign >95% of the population to a household bubble. These bubbles are exclusive and remain fixed throughout the simulation from May 11th onward.

We assume households in a social bubble to be fully connected 4 days out of 7 (i.e., the contact probability between any two bubble members per day is 4/7 = 0.57). We also test a higher and lower level of connectivity in terms of 7/7 and 2/7 days per week, respectively. Social contacts in a household bubble are implemented as a substitute of leisure contacts in the community and can be seen as repetitive leisure contacts with the same individuals. Therefore, the community contacts are reduced in proportion to the household bubble mixing to keep the overall contact rate unchanged. We also test household bubbles consisting of 3 and 4 households, where the number of household bubble contacts exceeds the number of simulated community contacts in our scenarios, so the total number of contacts increased. Symptomatic individuals have no social contacts with members of other households within their household bubble.

### Contact tracing strategy (CTS)

We implement contact tracing strategies (CTS) to assess their impact on hospital admissions if 70% of the symptomatic cases are considered to be index cases. Each index case is placed in home-isolation one day after symptom onset. One day later, unique contacts are traced and tested at a success rate of 90% for household members, and 70% for non-household members. We assume a false negative predictive value of 10%, as a combined outcome of sampling, lab-testing and clinical assessment of the treating physician. We performed sensitivity analyses regarding the proportion of symptomatic cases included as an index case, the false negative predictive value, the success rate to reach (non-)household contacts and contact tracing delays. The effect of these CTS parameters is tested using one of the social mixing assumptions (as described in the next paragraph).

### Scenario analyses

We defined different strategies by location-specific deconfinement strategies with structural uncertainty about social contact behavior after a lockdown. As such, we incorporated 4 mixing assumptions in our baseline scenario to capture a low and moderate increase in social contacts related to B2B and community activities. By modelling reductions in social mixing, we implicitly assume people either make fewer contacts compared to the pre-pandemic situation or the contacts they make are less likely to lead to transmission. For example, some transmission will be prevented by more frequent hand washing, distancing or the use of masks [22]. If we assume that social mixing at workplaces increased from 25% to 50%, we estimate the impact of “what if the risk of acquiring infection at work doubles compared to during lockdown, but remains still 50% less compared to pre-pandemic times”. Table 1 presents the social mixing details for each scenario.

**Table 1:**
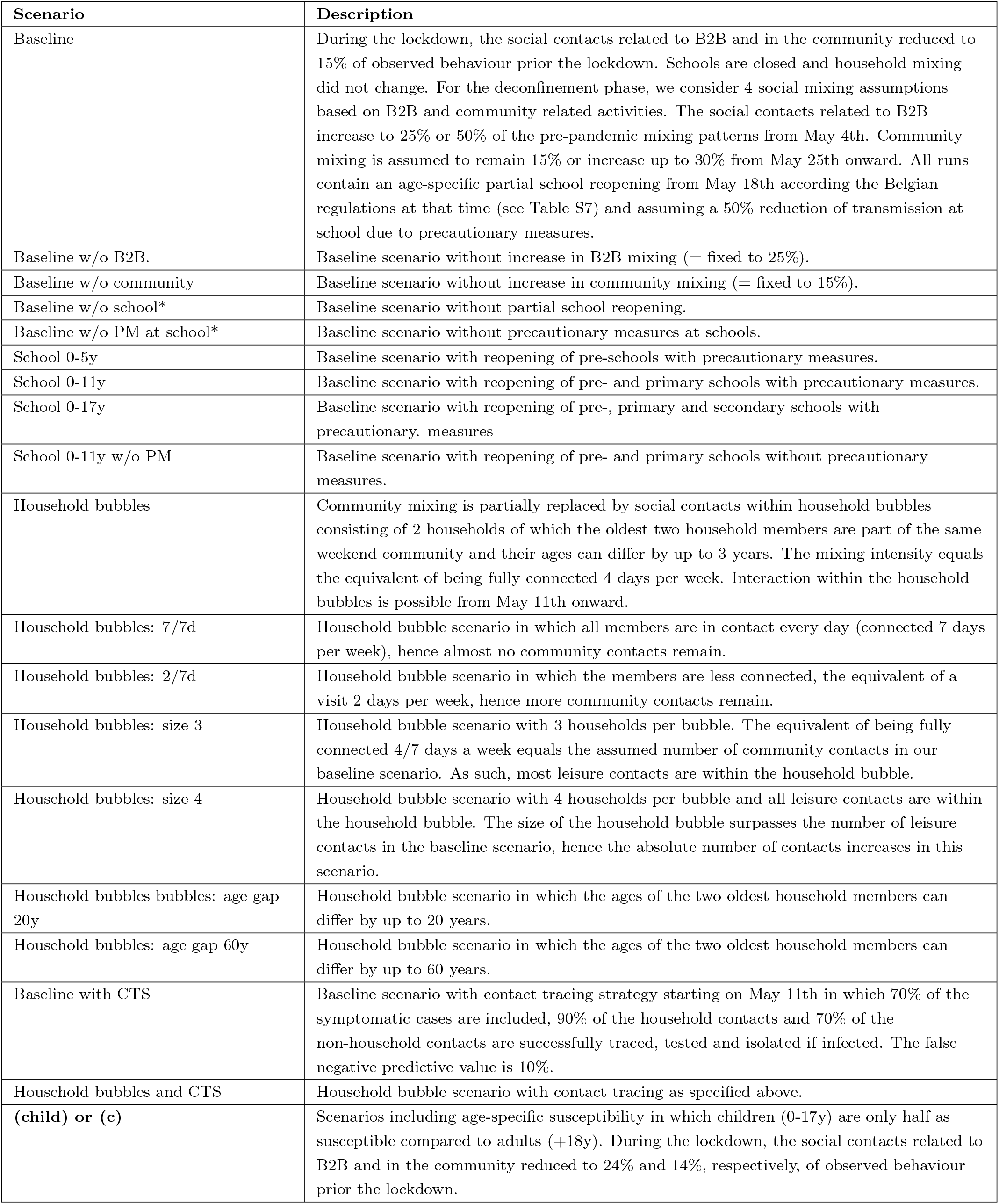
Scenario definitions. All reductions in social mixing are relative to observed social contact patterns before the lockdown. B2B: business-to-business, CTS: contact tracing strategy, w/o: without, PM: precautionary measures.

In our baseline scenario, we accounted for an increase of B2B mixing (i.e., contacts while at work) from May 4th up to 50% of the pre-pandemic observations. Business-to-consumer (B2C) and leisure transmission is harder to single out using social contact data within our model structure. To model the relaunch of economic activities and other (leisure) activities in the community, we incorporated a limited increase of community mixing up to 30% in our scenario analyses starting from May 25th. Note that we do not claim that the increase of community mixing is estimated to be 30% or restricted to this level, but we provide insights up to 30%.

For schools, we assumed a 50% reduction of transmission due to precautionary measures (smaller class groups, class separation, increased hand hygiene, etc.) and performed sensitivity analyses to explore the effect of these measures. We aligned the baseline scenario with the school regulations and timings for Belgium (see Table S7). In addition, we also included more general scenarios for re-opening pre-, primary and secondary schools from May 18th onward to make our analysis more explorative. We model that all schools close on July 1st, in line with the start of the national summer holiday period (until August 31st).

The Belgian government further relaxed restrictions in May 2020 by allowing additional contacts within the household context. We adopted a strict approach using household bubbles of two households of a similar generation based on the age of the oldest household member. To align a combined approach of household bubbles and contact tracing, both strategies start in our simulations on May 11th. We did not include additional region-specific distancing measures.

### Age-specific susceptibility

To fully explore age-specific effects, especially for school-related scenarios, we additionally calibrated our transmission model assuming that children (0-17y) are only half as susceptible compared to adults (+18y) [23]. The methods are provided in Supplementary Material.

### Sensitivity and robustness analyses

During the parameter estimation, we identified an ensemble of parameter sets at the intersection of the best scoring model runs according to the observed hospital admissions, doubling time before the lockdown and serial seroprevalence. The results presented in the main text are based on the single best parameter set, but we repeated the main scenarios (baseline, household bubbles, CTS and the combination of both) with the final ensemble of model parameters. To validate our choice for presenting results based on 10 stochastic realisations, we also ran our main scenarios with 20, 40 and 80 stochastic realisations.

## Results

We calibrated the transmission model up to April 30th, 2020, and continued all simulations up to August 31st to assess the impact of different deconfinement strategies. We start from a baseline scenario with step wise re-opening of B2B, schools and community activities including 4 assumptions capturing low and moderate increases in social mixing. Figure 1 presents the simulated hospital admissions over time from our baseline scenario with the timing of context-specific re-openings. Each gray line represents one stochastic trajectory of the simulator based on one social mixing assumption. The trajectories marked with A and B include an increase in community related social mixing, which has a clear impact on the projected hospital admissions. The trajectories marked with A and C include an increase in B2B related mixing. Without an increase in community mixing, the effect of B2B seems minimal. We estimated the reproduction number before the lockdown to be 3.42 [3.41-3.49], which dropped below 1 during the lockdown. The reproduction number in our baseline scenario increases above 1 after the deconfinement for community contacts, which includes B2C and leisure activities.

**Figure 1:**
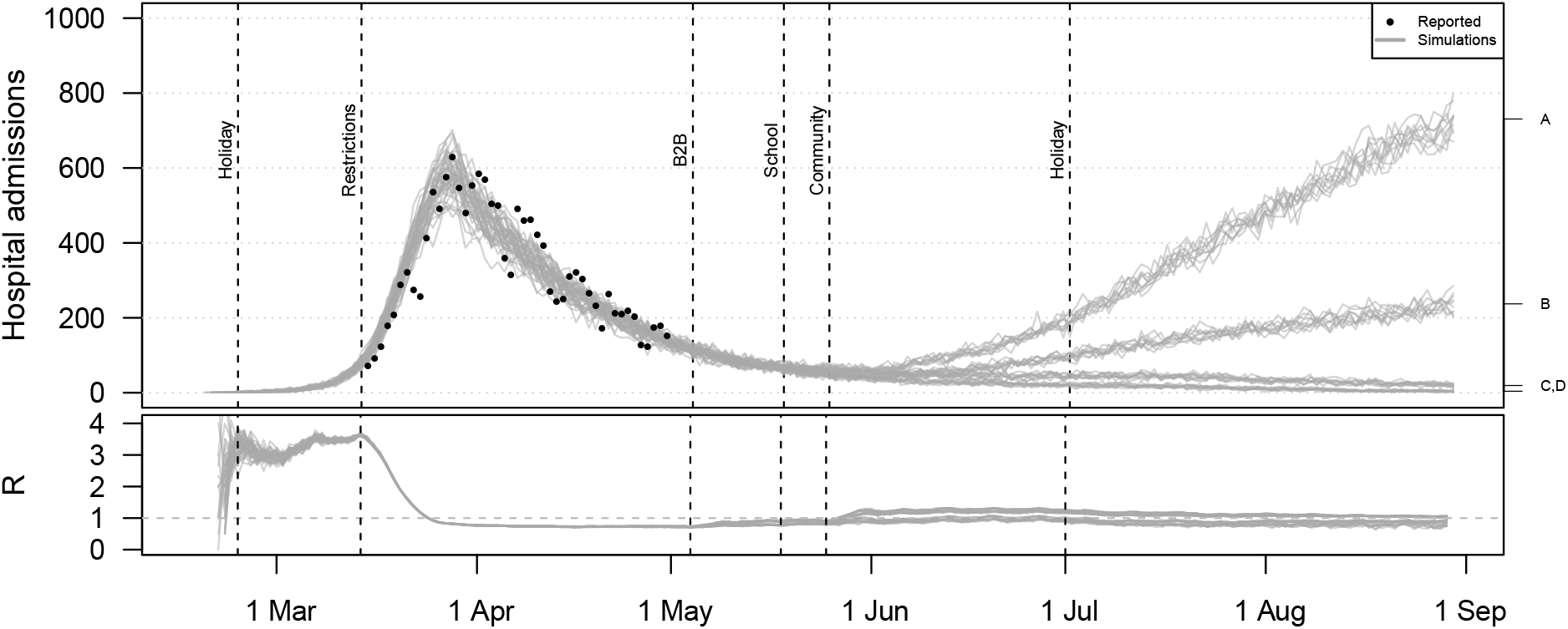
Hospital admissions and effective reproduction number (R) from the baseline scenario including 4 mixing assumptions. All simulations include social restrictions from March 14th and the partial school reopening in May. For the B2B, the social mixing after the lockdown is assumed to double from the indicated point in time (indicated on the right hand side with A and C) or to remain constant (B,D). Social mixing in the community is assumed to double (A,B) or to remain constant (C,D).

Scenario analysis shows that social mixing in household bubbles, contact tracing and a combined strategy has a clear impact on the hospital admissions over time (Figure 2). All scenarios are based on the same assumptions in terms of the absolute number of social contacts in line with the baseline scenario (Figure 1). If people have fewer unique contacts, as in the scenario that considers household bubbles, the number of hospital admissions decreases. This is also the case with a strict follow-up of symptomatic cases and their contacts when applying the contact tracing strategy. For both the household bubble and CTS scenario, the reduction is not sufficient if both B2B and community mixing doubles (trajectories marked with “A”), since the number of hospital admissions still increases over time. The combination of both strategies show a stabilising effect for all social mixing assumptions under study.

**Figure 2:**
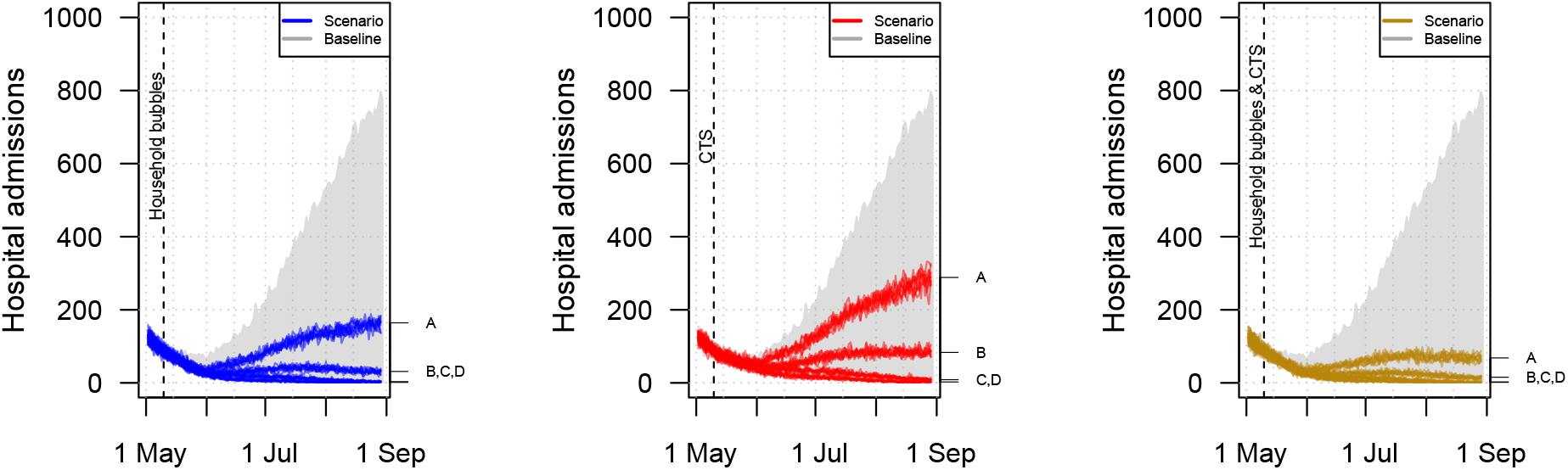
Hospital admissions over time when community mixing occurs in household bubbles (left), a contact tracing strategy (CTS) is in place (center), or both (right). All scenarios are based on the same natural disease history and quantitative mixing assumptions but differ from the baseline in terms of the network structure and application of contact tracing from the given point in time. The mixing assumptions A,B,C,D are explained in the caption of Figure 1.

Household bubbles are defined by connectivity, intergenerational mixing and size, which all have impact on our simulated hospital admissions, as presented by the projections for June and August in Figure 3. Note that the reported distribution and summary statistics strongly depend on our mixing assumptions so they can only be used to show relative differences across scenarios. Our default scenario with household bubbles shows an average reduction in the average number of hospital admissions by 53% in June and by 75% in August as compared to the baseline scenario.

**Figure 3:**
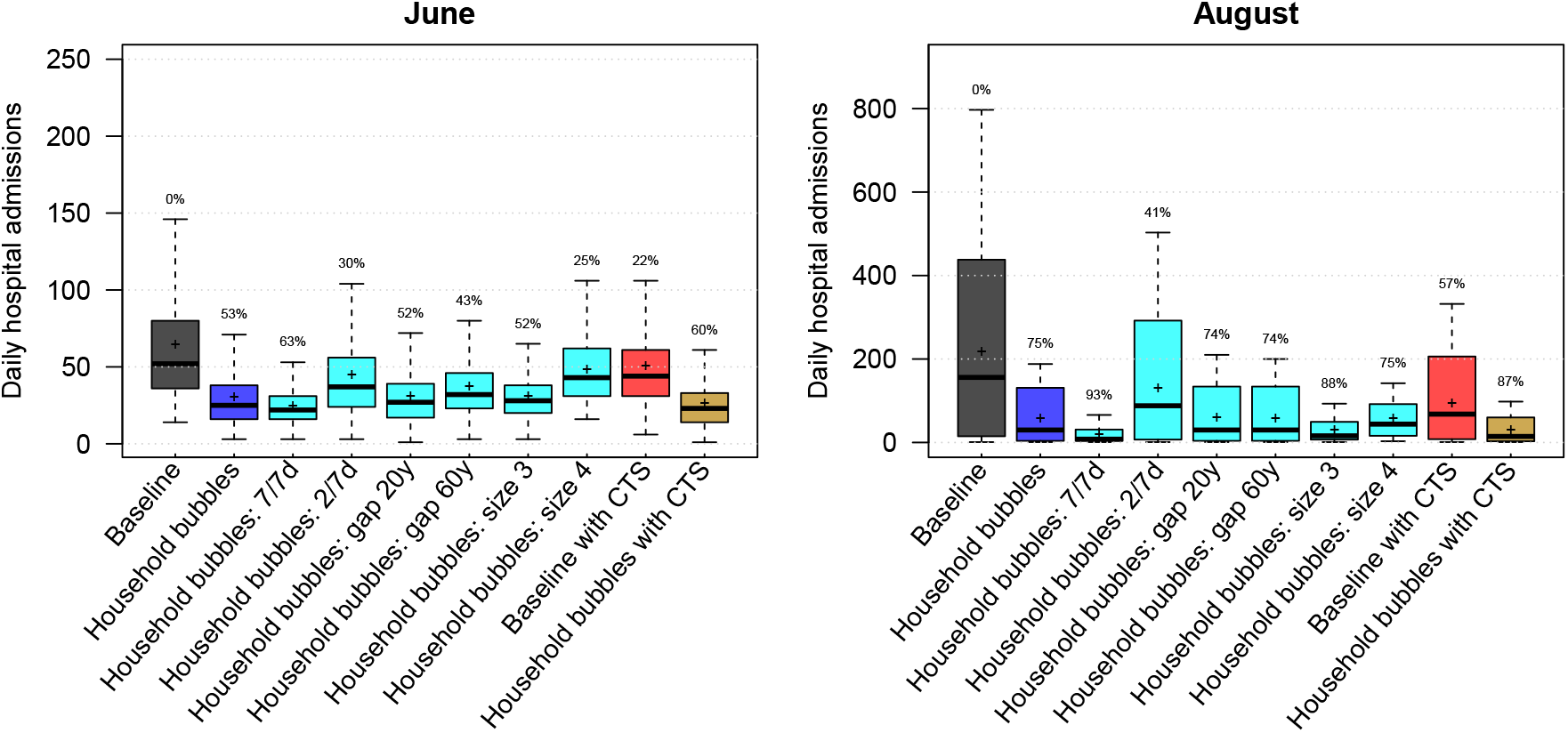
Distribution of the daily hospital admissions by June and August per scenario. The results are presented as the median (line), quartiles (box), 2.5 and 97.5 percentiles (whiskers) and average (cross) of the scenario results including social mixing uncertainty and stochastic effects. The percentage on top of the whiskers indicates relative reduction of the scenario average with respect to the baseline. CTS: contact tracing strategy.

By not having leisure contacts outside the household bubble (i.e. if the household bubbles are fully connected 7 days a week), the average number of hospital admissions can be reduced by 93% by August. If household bubbles are less strict (i.e. fully connected 2 days a week), the effect is less pronounced but the average number of hospitalizations in August can still be 41% less compared to our baseline. If household bubbles consist of households of which the ages of the oldest household members can differ up to 20 or 60 years and multiple generations are allowed within one household bubble, the effectiveness of this strategy decreases. The reduction in daily hospital admission by June is only 43% if 60-year differences are allowed. If household bubbles consist of 3 households, they almost replace all community contacts in our simulations, which results in fewer hospital admissions in the long run due to the closed network topology. If people mix within household bubbles of size 4, the average number of leisure contacts increases compared to our baseline mixing assumptions, which explains the reduced effectiveness of the household bubble approach for June. However, due to the closed nature of these extended bubbles and restricted number of unique contacts, the average number of hospital admissions by August is comparable to situation with household bubbles of size 2, despite the increased contact rates.

### Contact tracing

The follow up of symptomatic cases by strict isolation and contact tracing, i.e. testing of their contacts with isolation if infected, shows a substantial effect on the average number of hospital admissions (Figure 2 and 3). We project an average reduction in hospital admissions of 22% in June and 57% in August with the CTS in place, assuming that 70% of the symptomatic cases are subjected to contact tracing and comply with home isolation. The combination of contact tracing and repetitive social mixing in household bubbles has the potential to reduce the average number of hospital admissions up to 87% by August. This approaches the effect of strict household bubbles, but clearly allows more freedom in terms of social mixing.

Our CTS results are based on different assumptions with respect to timing and success rates of tracing, testing, and compliance to home isolation if infected. We performed a sensitivity analysis to challenge our CTS assumptions that 70% of the symptomatic cases are considered as index case, 10% false negative tests, a 90% success rate for tracing household members and 70% for other contacts. The false negative predictive value of testing, due to the sampling, lab-testing and assessment of the treating physician, is important but we still observed an impact of the CTS with 30% false negative tests if the coverage is high enough (see Figure 4). By varying the success rate of contact tracing per index case, the relative number of hospital admissions ranged from 35% to 60% of the base case scenario without CTS. Tracing non-household contacts seems to have the most impact, since their absolute number can be higher compared to household contacts. However, tracing and testing household contacts, which are easier to define and accessible via the index case, also makes a difference. With a maximum delay between symptom onset of the index case and isolation of infected contacts up to 4 days, we observed the best results in terms of averted hospital admissions. If this delay increases, the efficiency of CTS drops. Note that if index cases are identified and isolated only 6 days after symptom onset, the tracing should be very fast or there will not be much left to gain.

**Figure 4:**
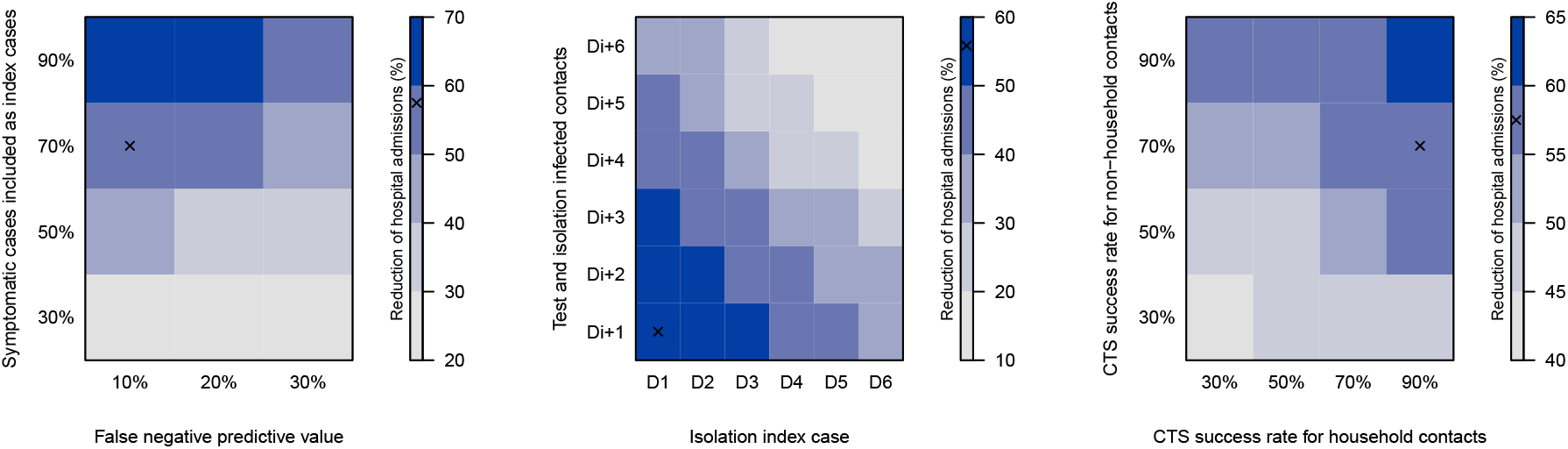
Reduction of hospital admissions due to contact tracing according to the symptomatic cases included as an index case, the false negative predictive value of testing, delays and the success rate of tracing, testing and isolating household and (non-)household contacts. Timings are expressed relative to symptom onset of index case (D0), and days after testing the index case (e.g., Di+2). All simulations start from the baseline scenario and assume a 50% and 30% reduction of B2B and community contacts, respectively, compared to pre-lockdown observations. The ‘x’ marks the default settings, which are used if a parameter is not shown. CST: contact tracing strategy

### Location-specific re-opening

We analysed the effect of location-specific deconfinement strategies on the total number of hospital admissions between May and August with two assumptions on the susceptibility for children up to 17 years of age: equally susceptible or only half as susceptible compared to adults (+18y) [23]. The results are presented in Figure 5. Starting from the baseline and each time leaving one location-specific re-opening out, we observed most impact of community mixing for both susceptibility-related assumptions. Without an increase in community mixing, the simulated hospital admissions decrease by almost 70%. Without an increase in B2B related social mixing, the total number of hospital admissions between May and August is still 50% less compared to our baseline scenario. The effect of household bubbles and CTS on the cumulative hospital cases is similar for both susceptibility assumptions. As expected, the impact of school re-opening is strongly associated with the assumption on age-specific susceptibility. Assuming that children are equally susceptible compared to adults, we observe an increase in hospital admissions up to 96% and 181% compared to our baseline scenario if schools re-open up to primary or secondary education, respectively. Re-opening primary schools without any precautionary measure such as smaller class groups, class separation, masks, and increased hand hygiene seems worse than opening all schools with a 50% reduction of transmission. The re-opening of pre-schools has limited effect on the simulated hospital admissions according to the mixing assumptions under study.

**Figure 5:**
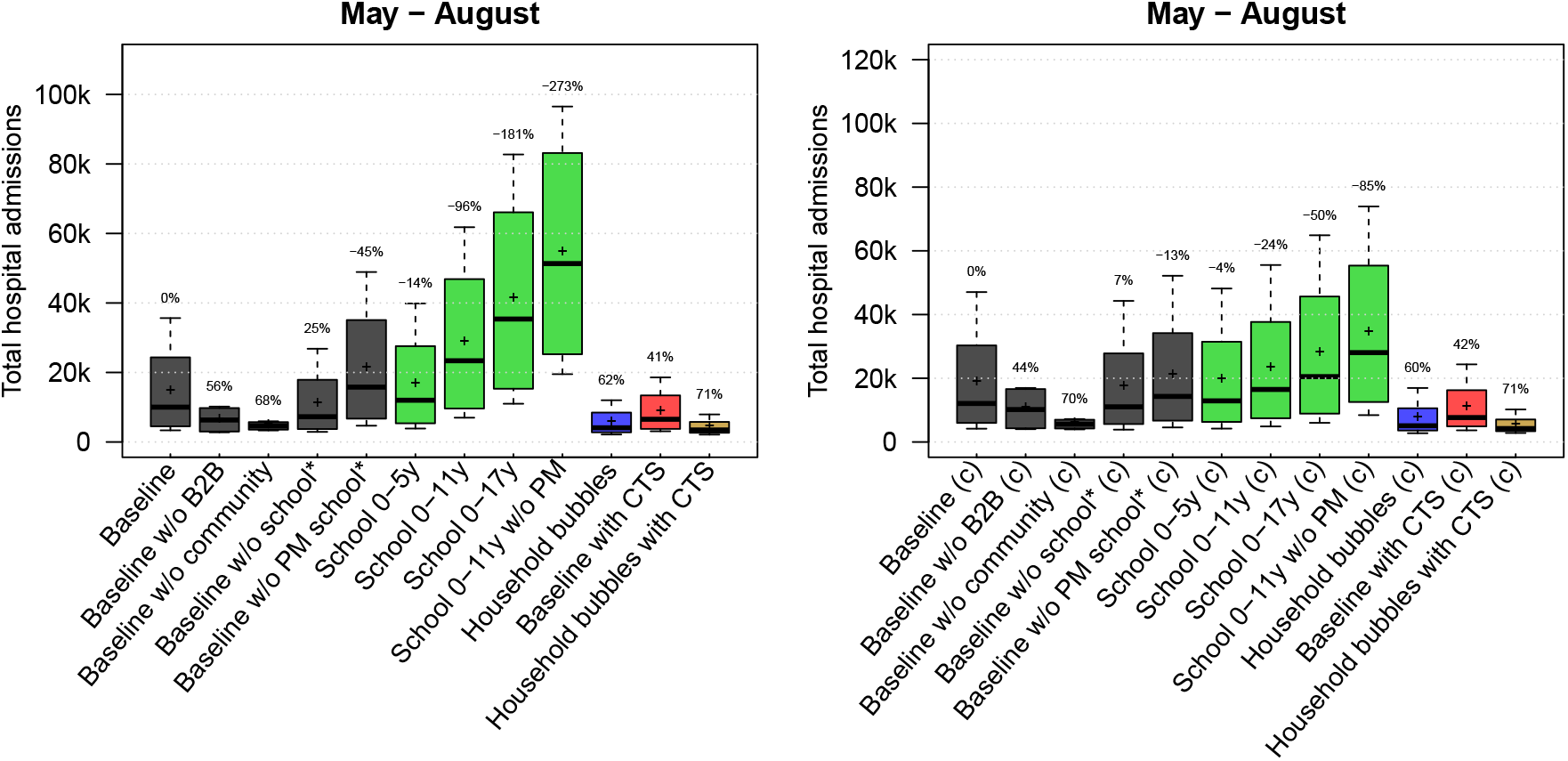
Total hospital admissions per scenario from May to August assuming that children (0-17y) are equally susceptible as adults (left) or only half as susceptible compared to adults (right). The results are presented as the median (line), quartiles (box), 2.5 and 97.5 percentiles (whiskers) and average (cross) for all combinations of the contact reductions per scenario. The percentages on top of the whiskers indicate the average reduction in hospital admissions with respect to the baseline. CTS: contact tracing strategy; w/o: without; PM: precautionary measures at school.

Assuming an age-specific susceptibility, re-opening primary schools has less impact on the predicted number of hospital admissions. If all children up to 17y of age go back to school with precautionary measures, we observed an increase of hospital cases up to 50% relative to our baseline scenario. Note that these scenarios do not take contact tracing or other physical distancing measures into account but express the transmission potential at school. Combining different scenarios to define the required contact tracing efficiency or other measures to enable schools to reopen is subject of future research.

### Sensitivity and robustness analyses

Given the correlated nature of our model parameters, different combinations can give a similar fit for the first wave but might lead to different outcomes for the deconfinement strategies in the scenario analyses. To assess the robustness of our results, we simulated the main scenarios with an ensemble of model parameter sets. The resulting projections in terms of hospital admissions over time (Figure S20) show more variation but the average reduction in hospital admissions (Figure S21) does not change.

We performed a robustness analysis on the number of stochastic realisations for the main scenarios and observe more spikes in the hospital admissions over time with an increasing number of stochastic realisations (Figure S16 and S17) but no differences in the average hospital admissions for June and August (Figure S18). Details are provided in the Supplementary Material S10 and S11.

## Discussion

Uncertainty on social mixing after a lockdown plays a crucial role in predicting the outcome of deconfinement strategies. How will people behave socially if restrictions are relieved? A deconfinement strategy can allow for economic or leisure activities, but people might still limit their contacts or they might fully exploit the renewed freedom, beyond what is requested but hard to regulate or enforce. In addition to contact frequency, contact intensity (duration, intimacy, indoor/outside location, etc.) also plays a role in the transmission dynamics. To handle this structural uncertainty in our simulations, we included different social mixing assumptions as part of assessing deconfinement strategies. Our baseline strategy including the 4 different mixing assumptions is not chosen to capture the observed situation as much as possible, but to analyse the relative impact of mutually exclusive scenarios as in comparative effectiveness research.

Parallel modelling work for the UK [24] showed that social bubbles reduced cases and fatalities by 17% compared to an unclustered increase of contacts. Social bubbles may be very effective if targeted towards small isolated households with the greatest need for additional social interactions and support. Their analyses confirm that social bubble strategy is an effective way to expand contacts while limiting the risk of a resurgence of cases.

We found a great potential for CTS to reduce transmission and hospital admissions, but it might not be enough to control future waves. The simulations in Figure 2 with the least strict physical distancing still show an increase in hospital admissions with CTS in place. Only if the number of contacts is limited and/or contacts take place in closed networks such as the household bubbles, the CTS is sufficient to keep the hospital admissions low. The relative proportion of symptomatic cases that is included in the CTS as index case is driving the efficiency. Also, timing is of the essence and contact tracing should start at the latest 4 days after symptom onset of the index case. The short serial interval makes it difficult to trace contacts due to the rapid turnover of case generations [25]. Keeling et al. [1] concluded that rapid and effective contact tracing can be highly effective in the early control of COVID-19, but places substantial demands on the local public-health authorities. We did not include or analyse the enhancing/spiraling effect when infected contacts are subsequently included as an index case. This could be one way to improve case finding to include as index case, which we implicitly incorporated in our strategy. Another effect of this spiraling approach might be a reduction of the workload given overlapping contacts with a previous index case. However, the timing of physical contacts and testing might interfere with this optimization procedure. We did not look into this but focus on the basic principles and stress the potential of CTS.

Kucharski et al. [26] also reported on the effectiveness of physical distancing, testing, and a CTS for COVID-19 in the UK. They concluded that the combination of a CTS with moderate physical distancing measures is likely to achieve control. They also used an IBM with location-specific mixing and transmission parameters and similar natural history of the disease. Their model is different in the number of contacts, which is fixed to 4, and social contact pools for school, work and other, are defined at a lower degree of granularity compared to our model. We are able to identify the class members and direct colleagues of infected individuals, and can confirm their conclusions on CTS and isolation strategies. We both stress the potential of CTS but warn that additional physical distancing measures are required to be successful.

Kretzschmar et al. [27] computed effective reproduction numbers with CTS and social distancing in place by considering various scenarios for isolation of index cases and tracing and quarantine of their contacts. Without a delay in testing and tracing and with full compliance, the effective reproduction number was reduced by 50%. With a testing delay of 4 days, even the most efficient CTS could not reach effective reproduction numbers below 1. We did not express the impact of CTS on the reproduction number, though also found a tipping point in the CTS effectiveness if contact tracing starts 4-5 days after symptom onset of the index case. To improve the early detection of cases, the use of universal testing in which the entire population is screened on a regular basis is promising [28].

School closure is considered a key intervention for epidemics of respiratory infections due to children’s higher contact rates [29, 30], but the impact of school closure depends on the role of children in transmission. Davies et al [23] conclude that interventions aimed at children might have a relatively small impact on reducing SARS-CoV-2 transmission, particularly if the transmissibility of subclinical infections is low. This is also the conclusion from our scenario analyses where we assume children (*<*18y) to be half as susceptible as adults (+18y).

Other IBM applications have been reported [31, 32, 3, 33] to simulate combinations of non-pharmaceutical interventions by targeting transmission in different settings, such as school closures and work-from-home policies. Modelling the isolation of cases in safe facilities away from susceptible family members or by quarantining all family members to prevent transmission has shown substantial impact. Models that explicitly include location-specific mixing are very relevant for studying the effectiveness of non-pharmaceutical interventions, as these are more dependent on community structure than e.g. with vaccination [31]. However, implementing the available evidence into a performant and tailor-made model that addresses a wide range of questions about a variety of strategies is challenging [32, 2].

Although our analysis is applied to Belgium, our findings have wider applicability. We considered the effect of universal adjustments in terms of social mixing (isolation, repetitive contacts, contact tracing). We modelled 11 million unique inhabitants with detailed social contact patterns by age and location. Hence, we can compare model results with absolute incidence numbers in the absence of premature herd immunity effects due to a reduced population size. The latter can be an issue for models that use a scaling factor to obtain final results. Our individual-based model provides a high-resolution, mechanistic explanation of the reproduction number and transmission dynamics that are relevant on a global scale.

### Limitations

Any model is a simplification of reality and therefore depends on the assumptions made. In addition, our spatially explicit IBM is calibrated on national hospitalization data so uncertainty is inevitably underestimated. As such, we rely on scenario analyses and further sensitivity analyses are necessary. Model results should therefore be interpreted with great caution. Our IBM is a mechanistic mathematical model that uses conversational contacts as a proxy of events during which transmission can occur. By definition, SARS-CoV-2 infection events that occurred through the environment (e.g., contaminated surfaces) are covered by these conversational contacts.

The use of antiviral drugs in combination with CTS can reduce the effect of local outbreaks [34]. This kind of pharmaceutical intervention is not incorporated in the current analysis. We focused on the transmission dynamics in the general population and did not consider care homes separately in our analysis. They form predominantly a sink for infections, with high morbidity and mortality, but are not likely to drive the transmission. To focus on the disease burden in the elderly, the social interactions within elderly homes and with their environment become more important [35]. We did not include aspects related to travel or weather conditions (UV light, humidity, temperature) which may impact both transmission and social contact behaviour in ways that are still largely unknown.

## Data Availability

We provide all code and data in an open-source GitHub repository: https://github.com/lwillem/stride_covid19_v1

## Abbreviations

STRIDE: Simulator for the TRansmission of Infectious DiseasEs
w/o: without
B2B: Business-to-Business
B2C: Business-to-consumers
ICU: Intensive Care Unit
CTS: Contact Tracing Strategy

## Authors’ contributions

LW, PB and NH conceived the study. SA, PJKL, PC, OP, SM, JW, SAH and CF contributed to the data collection and analysis. PJKL and EK contributed to the software development. All authors contributed to the final version of the paper and approved the final manuscript.

## Acknowledgements

The authors are very grateful for access to the data from the Belgian Scientific Institute for Public Health, Sciensano, and from the Vaccine & Infectious Disease Institute (VaxInfectio), University of Antwerp. We acknowledge support from the Antwerp Study Center for Infectious Diseases (ASCID) and Flemish Super-computer Centre (VSC) with special thanks to the CalcUA-team (FB and SB). We thank several researchers from the SIMID COVID-19 consortium from the University of Antwerp and Hasselt University for numerous constructive discussions and meetings.

## SUPPLEMENTARY MATERIAL

### S1 Model population

This work builds upon a stochastic individual-based simulator, STRIDE, we developed for influenza [1, 2] and measles [3]. Our individual-based model has a particular focus on social contact patterns by modelling each individual as part of “contact pools”, representing a household, school-class, workplace, or community.

Household combinations, which specify the age of each member, are based on Belgian census data from 2011. We had to process the census data to ensure anonymization by excluding households containing 7 or more individuals (3.5% of population) and by using age groups for household combinations with a frequency of less than five. As such, we aggregated ages into 2-year intervals for individuals aged 0-25 and 5-year intervals for individuals over 26 years of age. If the frequency of an aggregated household composition still remained less than five, the households were excluded (0.7% of population). Next, we re-sampled ages from the age intervals to settle each household combination for our model population. We matched our resulting household data with summary statistics for household size and noticed an under-representation of large households containing children. Therefore, we duplicated 25,000 and 45,000 randomly chosen households of size 5 and 6, respectively, in which the second youngest household member was of age 0-25. These numbers were chosen to obtain matching distributions regarding household size, age in the population and age per household size. Figures S1 and S2 present summary statistics from the model population and Belgian census data.

**Figure S1:**
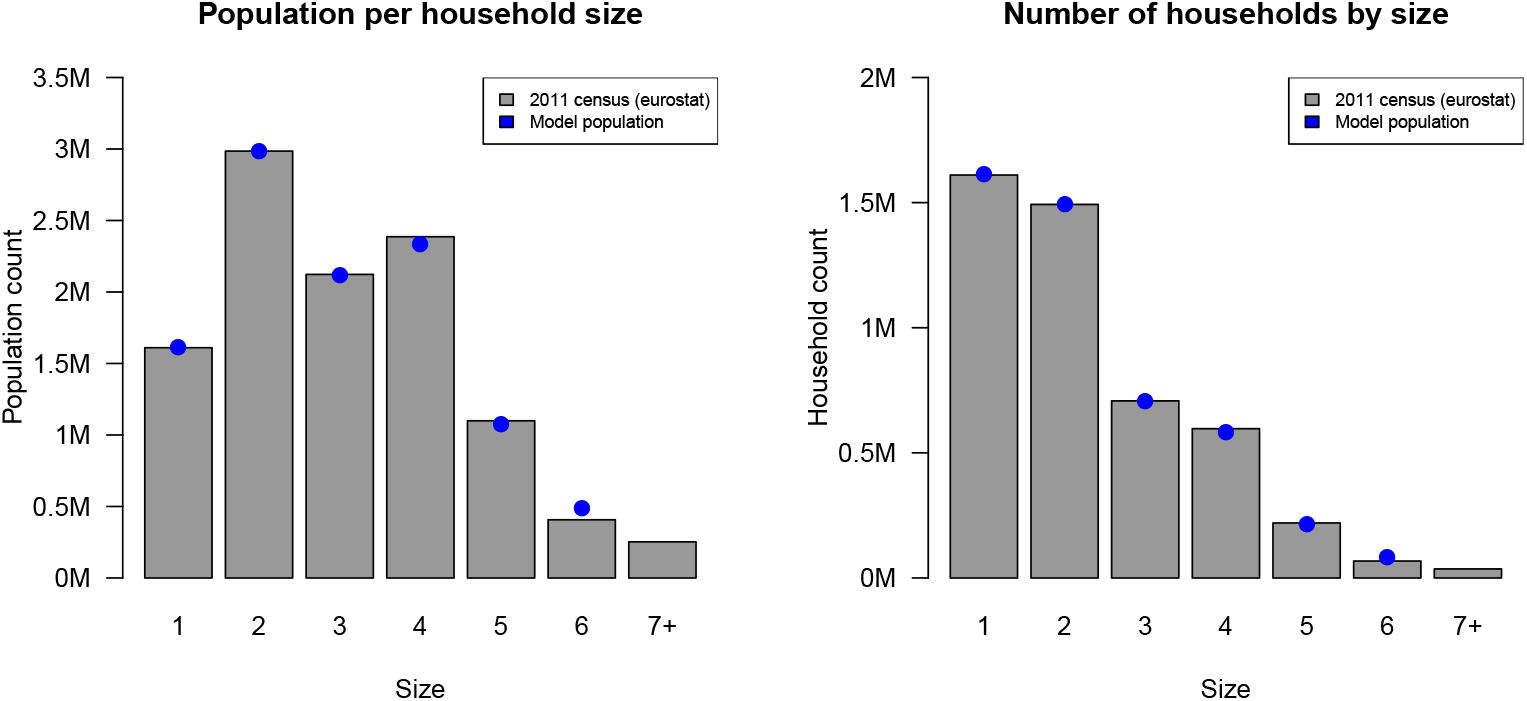
Population count per household size and number of households per size: Belgian 2011 census and model population. Numbers are expressed in million (M).

We build our population by sampling households and assigning them to geographic locations based on the population census of 2001. Our population of 11 million people is closed, meaning that no births or deaths occur during the simulation, nor emigration or immigration. Children are assigned to a daycare center (0–2 years old), pre-school (3–5 years old), primary (6–11 years old), secondary (12–17 years old) or tertiary (18–23 years old) school based on Belgian enrolment statistics from Eurostat [4]. Daycare centers in Belgium comprise on average 8 infants [5], with a skewed distribution up to 18 infants. School classes in pre-, primary and secondary schools contain on average 19, 20 and 20 children, respectively, using a class size distribution based on government statistics [6]. Students enrolled in tertiary schools are assigned to groups of 50 fellow students on average. Adults (18–64 years old) are assigned to a workplace or a daycare center/school class based on age-specific employment data and aggregated workplace size data from Eurostat [4]. We included one adult per 8 children in a daycare center and one adult per class in the pre- and primary school setting. Each “workplace” represents professional contacts in line with “business-to-business” (B2B) activities. The size of the workplaces is based on data from Eurostat and categorized into 1-9 (94%), 10-19 (3%), 20-49 (2%), 50-249 (0.8%) and +250 (0.2%) people. Geographic workplace assignment is based on commuting data from the Belgian 2001 census.

To represent leisure activities, family visits, “business-to-consumer” (B2C) and other contacts, the model contains “communities”. Each community is specified by a geographic center based on population density and contains on average 500 individuals. This arbitrary number affects contact probabilities but it does not influence contact rates. Each individual is assigned to one of three nearest community centers close to their home to represent weekday interactions and activities. For weekends, individuals can be assigned to the same community center or another one close to home. This community setup allows individuals to have similar contacts during week and weekend days, but prevents a strict compartmentalisation of the population.

**Figure S2:**
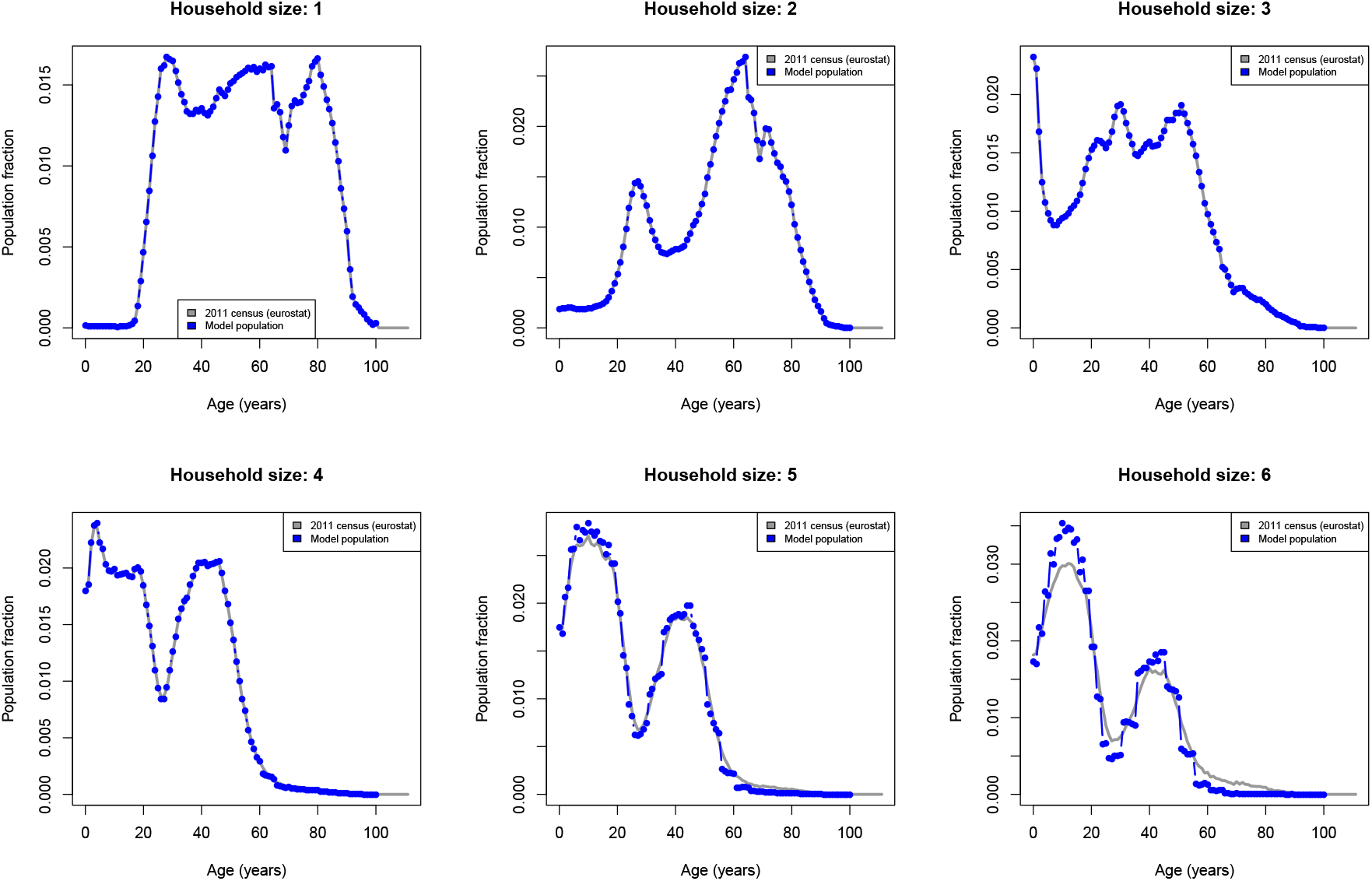
Age profile per household size from the Belgian 2011 census and the model population.

### S2 Social contact patterns

Social mixing in STRIDE is based on a diary-based social contact study performed in Belgium in 2010-2011 [7, 8, 9]. All participants were asked to record their contacts during one randomly assigned day. They also reported their time-use by activity, location and distance from home. Table S1 provides an overview on how the survey data is used to inform contact pools in the individual-based model. Note that contact rates at school and at work are conditional upon school enrolment and employment, respectively. During each time step, we match the age- and location-specific contact rate (= number of contacts per day) with the number of individuals in a contact pool (e.g., workplace, school, etc) to calculate the contact probability. We limit contact probabilities at 0.999, to prevent deterministic system behavior, and always use this maximum in a household setting since random mixing between household members is an adequate approximation of social contact behaviour for infections transmitted via close contacts [10]. Age-specific contact matrices are aggregated in the model into a vector containing age-dependent contact rates before calculating the contact probability. If people of different ages are part of the same contact pool, we use the lowest age-specific probability in their Bernoulli trial to enable a contact event.

The model allows to track social contacts in the population on a daily basis for all or for a random selection of individuals. The output is aligned with Socialmixr [11] and SOCRATES [12] to generate and visualise aggregated social contact matrices. Figure S3 presents social contact patterns from STRIDE summarized in 2×2 matrices based on 5000 individuals during weekend days and Figure S4 for weekdays before the COVID-19 lockdown.

**Table S1:** Implementation of “contact pools” in STRIDE based on social contact survey data [7, 8, 9]. Participants with more than 20 professional contacts per day (with students, clients, patients, etc.) had to report only the total number and age groups of their Supplementary Professional Contacts (SPC).

| Reported location in the social contact survey | Contact details in the social contact survey | Survey participant selection | Contact pool in STRIDE |
| --- | --- | --- | --- |
| Home | Household members | All | Household |
| Home | Non-household members | All | Community |
| School | All | “Student” (or age <18y) and time-use data contains “school” | School |
| Work (B2B) | Adults (>17y), >15min and non-SPC | “Employed” and time-use data contains “work” | Workplace |
| Work (Teaching) | Children (0-11y) | “Employed” and time-use data contains “work” | School |
| Work (B2C and teaching) | non-B2B contacts | “Employed” and time-use data contains “work” | Community |
| Leisure, transport, family, grandparents, other | All | All | Community |

**Figure S3:**
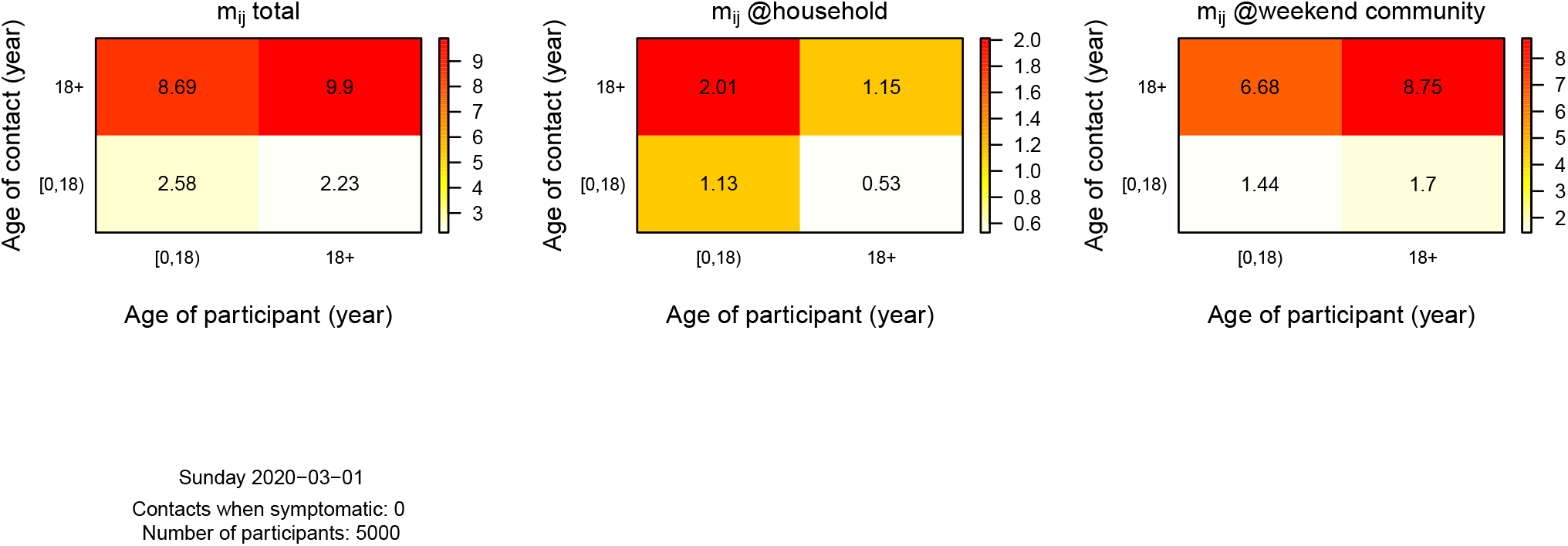
Social contact matrices based on 5000 individuals within STRIDE during weekends. These are aggregated 2×2 matrices based on one-year age group data from the model.

**Figure S4:**
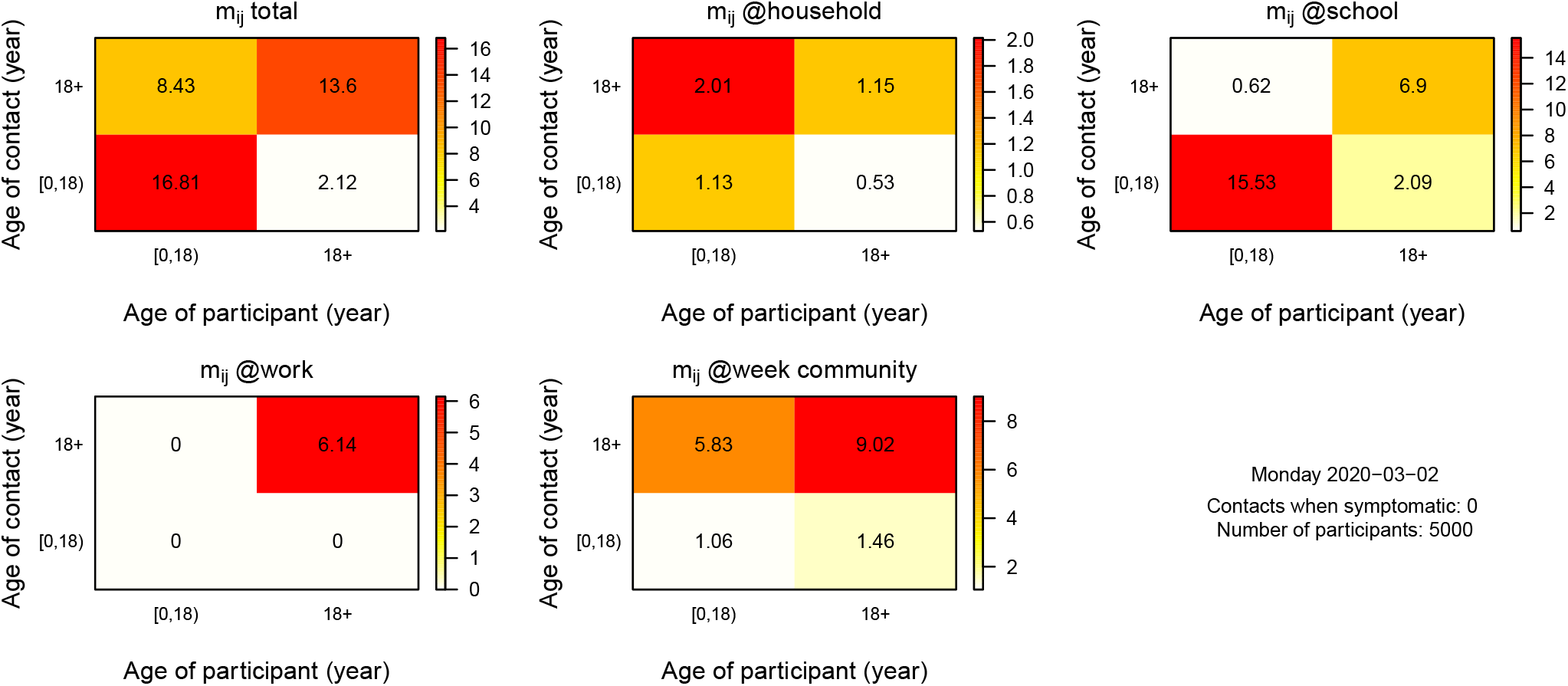
Social contact matrices based on 5000 individuals within STRIDE during weekdays. These are aggregated 2×2 matrices based on one-year age group data from the model.

### S3 Age-specific probability to be symptomatic

We assumed an overall proportion of symptomatic cases in the population before lockdown measures of 50% based on Li et al. [13]. To obtain an age-specific probability of symptomatic cases, we combined this population estimate with the age-specific relative susceptibility to symptomatic infection reported by Wu et al. [14]. Figure S5 presents the age-specific data from [14], which we amended with an assumption for individuals 0-19 years of age. These relative proportions had to be re-scaled and weighted by age to end up with a population average of 50%. Therefore, we calculated the relative population size by age as:

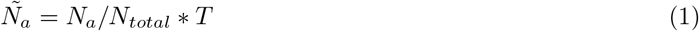

with *N*_*a*_ the population size of age group *a, N*_*total*_ the total population size and *T* the number of age groups. Secondly, we calculated the age-specific probability to be symptomatic as:

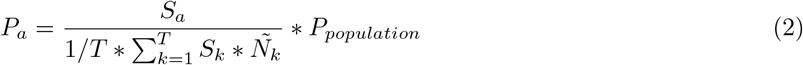

with *S*_*a*_ the relative susceptibility to symptomatic infection for age *a, N*^∼^_*a*_the relative population size for age *a*, and *P*_*population*_ the proportion symptomatic cases on the population level. Figure S5 presents the resulting probabilities to be symptomatic by age. Note that we had to truncate the highest relative susceptibility to symptomatic infection to maintain all age-specific probabilities between 0 and 1. This limitation is due to the interaction between the age-specific susceptibility and population sizes with the overall proportion of 50%.

The proportion of symptomatic cases per age group is disease-related and fixed over time. The proportion of symptomatic cases in the population depends on the age of the newly infected cases, which is driven by social contact and transmission dynamics. Given the temporal aspects of social contact behavior and restrictions, the overall proportion of symptomatic cases in the population can change over time.

**Figure S5:**
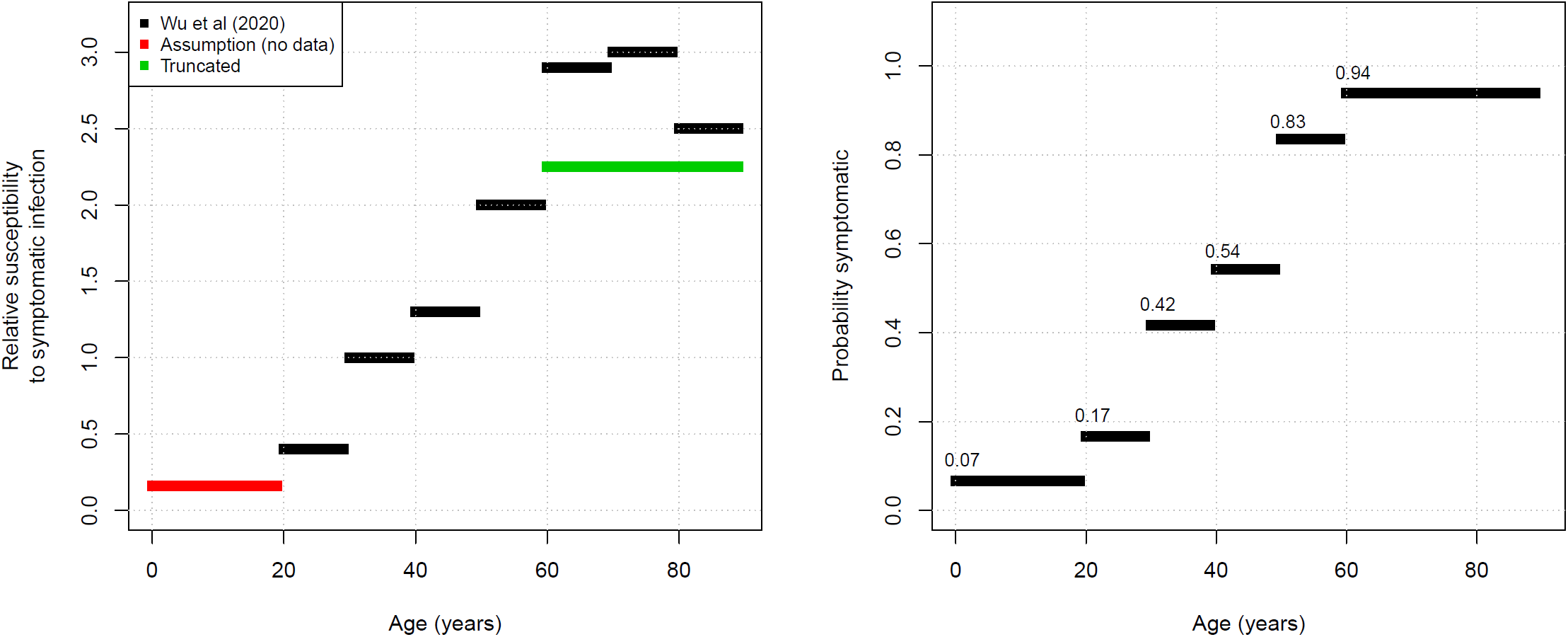
Relative susceptibility to symptomatic infection by age based on Wu et al. [14] (left) and estimated probability of symptomatic infections (right).

### S4 Natural disease history

The individual-based model STRIDE for COVID-19 is based on susceptible, exposed, infectious and recovered individuals, with the infectious health state divided in pre-symptomatic, symptomatic and asymptomatic. Throughout this text, we use “asymptomatic” for all cases that do not experience any symptoms throughout their infection. We assume that the latent and infectious period for asymptomatic cases is similar to symptomatic cases, so the estimations for the start and duration of the infectious period are generalized for asymptomatic cases. The level of infectiousness increases after symptom onset. Figure S6 presents the overall disease dynamics. Hospitalization is not part of the transmission model and added with a dashed line in combination with the age-specific rate based on a proportionality and delay factor.

**Figure S6:**
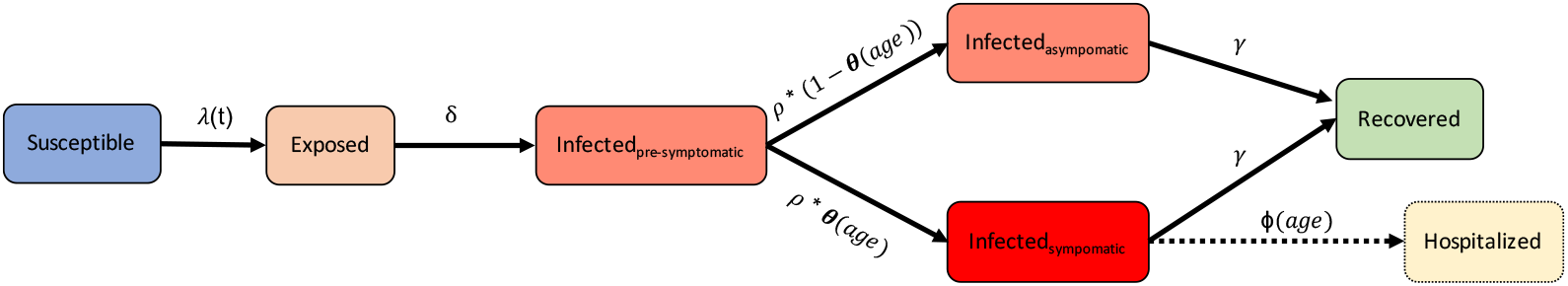
Overall disease dynamics. The infection process is driven by the force of infection *λ*(*t*), latent period 1*/δ*, pre-symptomatic period 1*/ρ*, proportion symptomatic *θ*(*age*), recovery rate *γ* and hospitalization rate *ϕ*(*age*).

From the individual perspective (see Figure S7), each infected case experiences an incubation period, an infectious period and optionally a symptomatic period. The start of the infectious period is related to the incubation period and this dependency is captured in our model with a “pre-symptomatic” infectious period rather than characterising the latent period. This modelling choice is driven by the literature on the incubation period and pre-symptomatic infectiousness [15, 13]. Hospital admissions are calculated post-hoc based on the model output. For each symptomatic case, there is a likelihood to be hospitalised and a delay distribution to specify the time between symptom onset and hospital admission. This likelihood and delay are age-specific and are described in Section S6.

**Figure S7:**
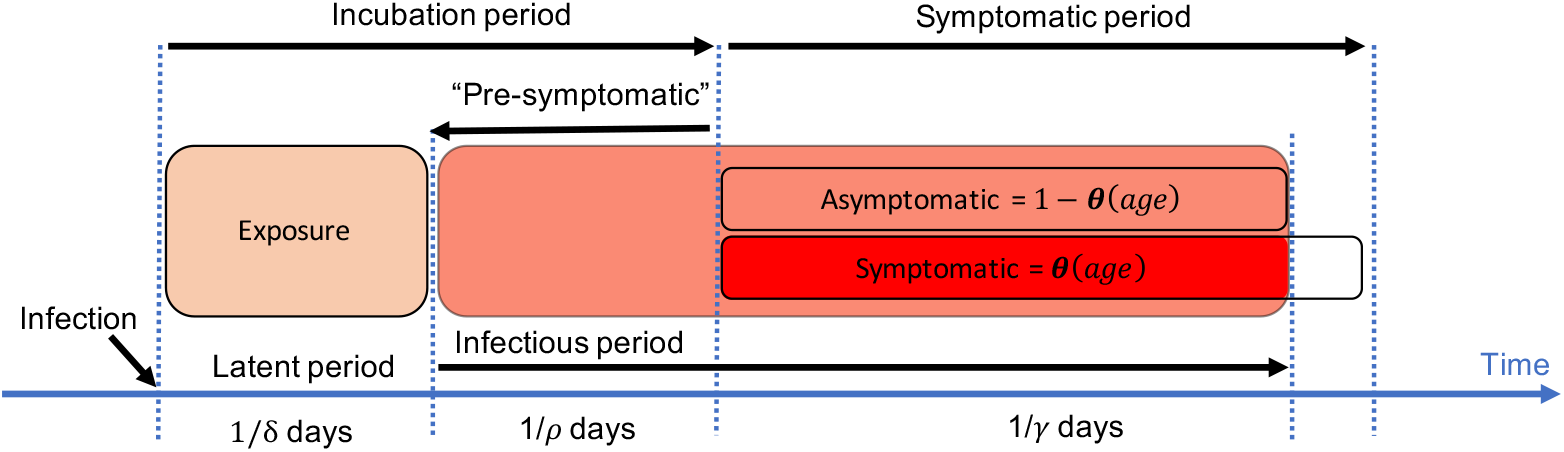
Disease dynamics in the STRIDE model on the level of the individual. Exposed individuals become infectious without symptoms after 1*/δ* days. Since not all cases develop symptoms 1*/ρ* days later, we denote this second stage as “pre-symptomatic”. After 1*/γ* days, individuals are not infectious anymore. Disease parameters are described in Table S2.

In practice, we define for each infected case an incubation period and whether the case is symptomatic or asymptomatic. Relative to the end of the incubation period, we specify the start of the infectious period and the duration. The following paragraphs describe how we derive the probability functions for the incubation, pre-symptomatic, infectious and symptomatic period.

The start of the infectious period, relative to symptom onset, is based on the results from He et al [15], who concluded that viral shedding may begin 5 to 6 days before the appearance of the first symptoms. They conclude that after symptom onset, viral loads decrease monotonically and decline significantly 8 days after symptom onset, as live virus could no longer be cultured. The inferred infectiousness profile was captured in a shifted gamma distribution with shape 20.52, rate 1.59 and shift 12.27 days. To estimate the pre-symptomatic infectious period density for STRIDE. We truncated the shifted gamma distribution at −1 by dividing the probability density function by the cumulative distribution function evaluated at −1 (left panel of Figure S8).

**Figure S8:**
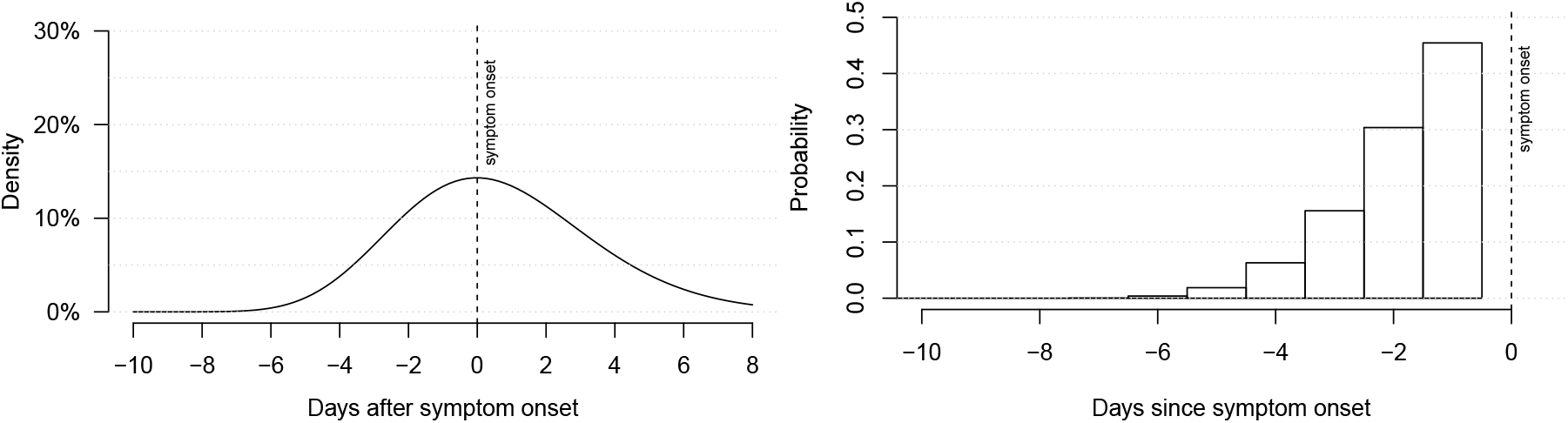
Infectiousness profile relative to symptom onset from He et al [15] (left) and the discrete pre-symptomatic infectious period distribution for STRIDE (right).

The incubation period in STRIDE is based on the reported mean of 5.2 days (95% confidence interval: 4.1 to 7.0) from Li et al. [13]. The distribution follows a log-normal distribution with logmean=1.43 and logsd=0.66. The numerical parameters are derived from He et al. [15] and are represented by the density plot in Figure S9. We calculated the discrete version of this log-normal distribution with the assumption that infectiousness has to start at least one day prior symptom onset and at least one day after infection. As a result, the incubation period is at least 2 days, which results in the probability distribution as presented in Figure S9;

**Figure S9:**
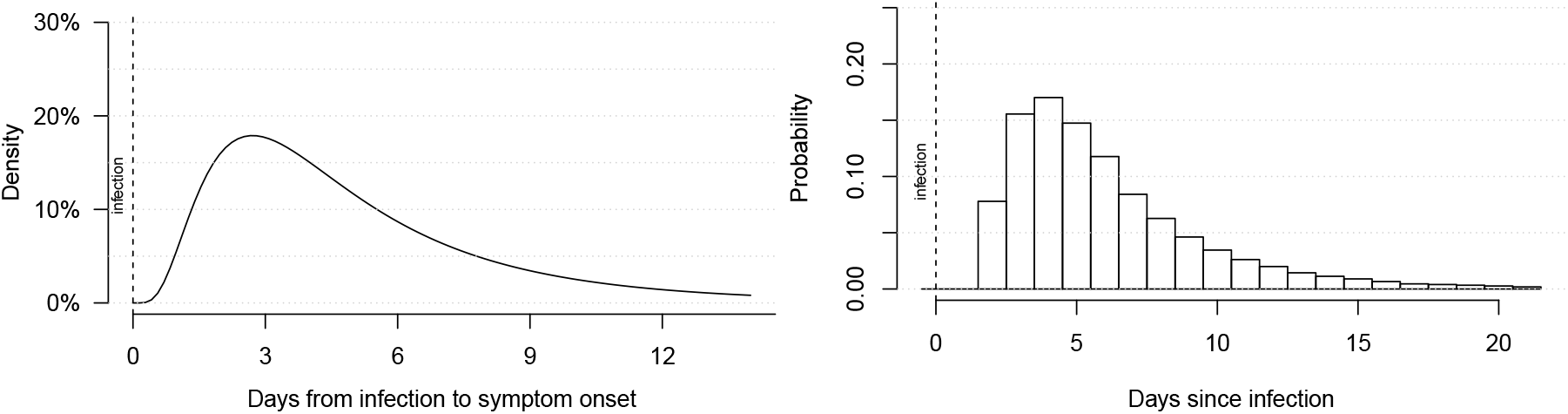
Incubation period distribution from Li et al. [13] (left) and the standardized discrete distribution for STRIDE (right).

He et al. [15] reports that infectiousness declines after symptom onset up to a maximum of 7 days, which restricts the overall infectious period. The modelling study by Lourenco et al. [16] used a normal distribution with a mean of 4.5 days and standard deviation of 1. Given their SIR model structure, this value can not be used directly, but was used as prior for our parameter estimation. We evaluated normal distributions with mean 5, 6 and 7 days and standard deviation 1 with the aim that the combination of the pre-symptomatic period and the total infectious period resembles the infectious profile relative to symptom onset from He et al.[15]. With a mean infectious period of 6 days, the number of individuals in STRIDE that are still infectious 7 and 8 days after symptom onset corresponds to 3% and *<*1%, respectively, which is in line with the results [15]. The infectious period distributions relative to infection and symptom onset are presented in Figure S10.

The focus of the transmission model lies on new infections over time and the behaviour of symptomatic cases after their infectious period has no impact on transmission dynamics. Therefore, we choose to fix the symptomatic period to the maximum infectious period after symptom onset to 7 days. The infectiousness of asymptomatic cases in STRIDE is set to 50% compared to symptomatic cases, based on Li et al. [13].

**Figure S10:**
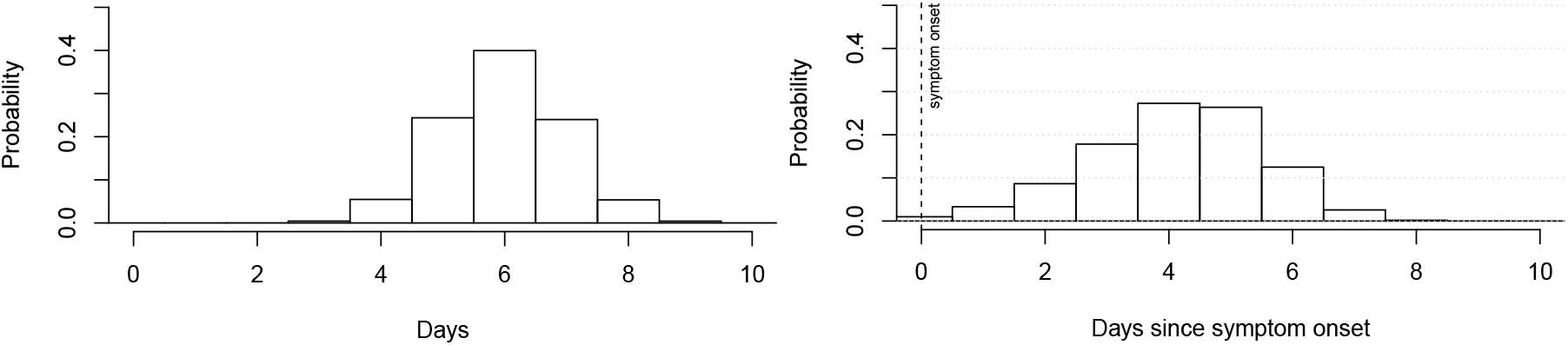
Infectious period distribution in STRIDE in total (left) and relative to symptom onset (right)

**Table S2:**
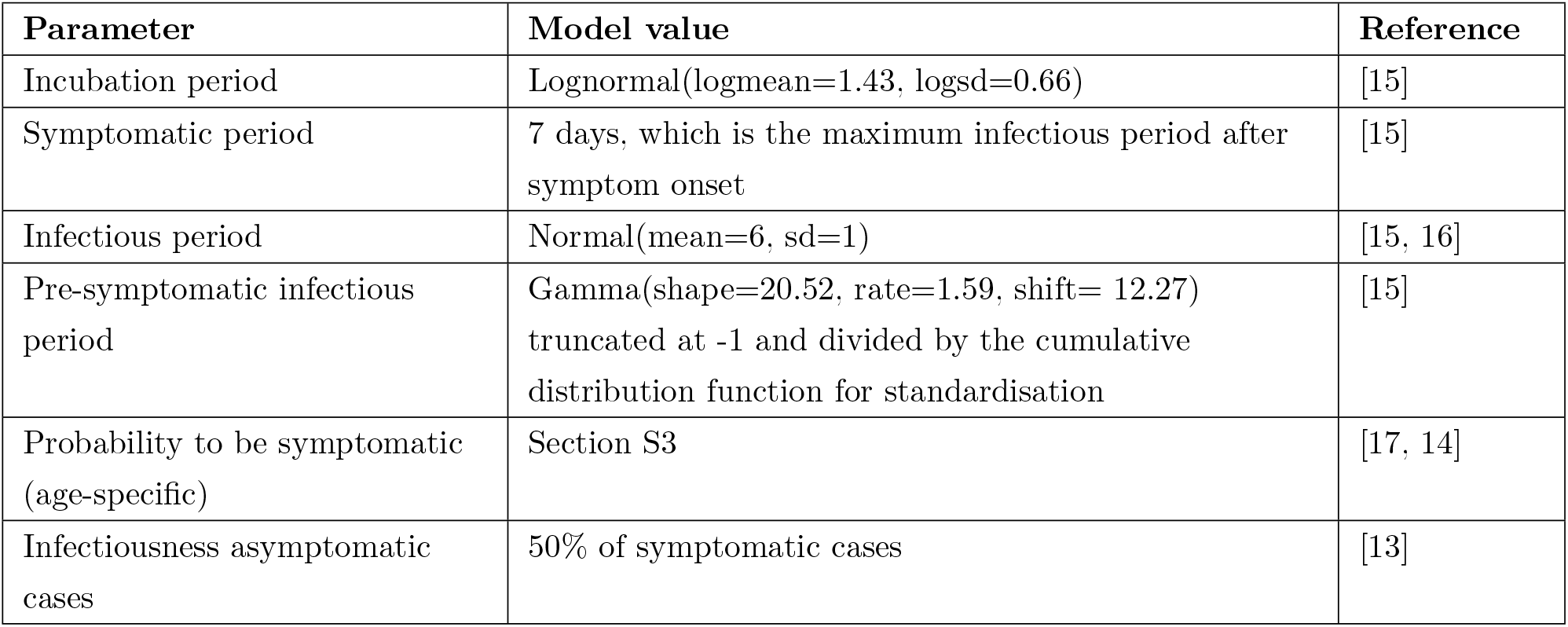
Summary table with disease characteristics. Transition parameters are discretized to the modelling time step of 1 day

| Parameter | Model value | Reference |
| --- | --- | --- |
| Incubation period | Lognormal(logmean=1.43, logsd=0.66) | [15] |
| Symptomatic period | 7 days, which is the maximum infectious period after symptom onset | [15] |
| Infectious period | Normal(mean=6, sd=1) | [15, 16] |
| Pre-symptomatic infectious period | Gamma(shape=20.52, rate=1.59, shift= 12.27) truncated at -1 and divided by the cumulative distribution function for standardisation | [15] |
| Probability to be symptomatic (age-specific) | Section S3 | [17, 14] |
| Infectiousness asymptomatic cases | 50% of symptomatic cases | [13] |

### S5 Transmission probability per contact

The basic reproduction number (R_0_) captures the combined effect of social contact patterns, demography, disease dynamics and the probability of transmission given contact. By keeping everything constant except the transmission probability, we can fit the relation with R_0_ by counting the number of secondary cases per index case in a susceptible population. The goal is to easily translate a transmission probability given contact to R_0_ and vice versa. Especially the latter is convenient to specify R_0_ as model parameter. As such, a given R_0_ is converted into a transmission parameter to be used in the simulation model. To enable this conversion, we need a continuous function for the number of secondary cases in terms of the transmission probability, which we can invert.

For the entire range of transmission probabilities, this relation needs to go through zero and levels off by the total number of unique contacts within the infectious period for the simulated population, social contact dynamics and disease characteristics. Capturing the full relation between the transmission probability and the secondary cases falls outside the scope of this research, since our goal is to simulate slight variations of the transmission probability to obtain R_0_ values in range with published values. As such, we focus on a specific range of the transmission probability and assume a linear relation between the transmission probability and the average number of secondary cases in a susceptible population. Based on preliminary runs, we defined a range for the transmission probability between 0.01 and 0.14, which corresponds with an average number of secondary cases between 1 and 5.

Each simulation with a specified transmission probability started with 20 randomly infected cases between 1 and 99 years of age, and traced their secondary cases. Note that newborns are never selected as index cases. The choice for 20 infected cases instead of 1 in a fully susceptible population according the definition of R_0_ is to reduce the number of model realisations with factor 20. On 11 million people, the difference between 1 and 20 infected seeds is inferior. To capture temporary effects for (pre-)symptomatic infectious periods during week and weekend days, we ran different simulations for each transmission probability starting from Monday the 1st up to Sunday the 7th of February 2020.

For each transmission probability (15x) and starting day (7x), we ran 10 stochastic realisations starting with 20 infected cases. As such, our fitting procedure to capture the relation between the transmission probability and the (basic) reproduction number is based on the secondary cases of 21000 index cases. Figure S11 presents the results of these simulations and the linear model we fitted through these data:

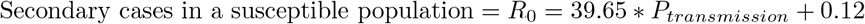

To use R_0_ as model input, we use the inverse:

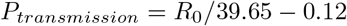

**Figure S11:**
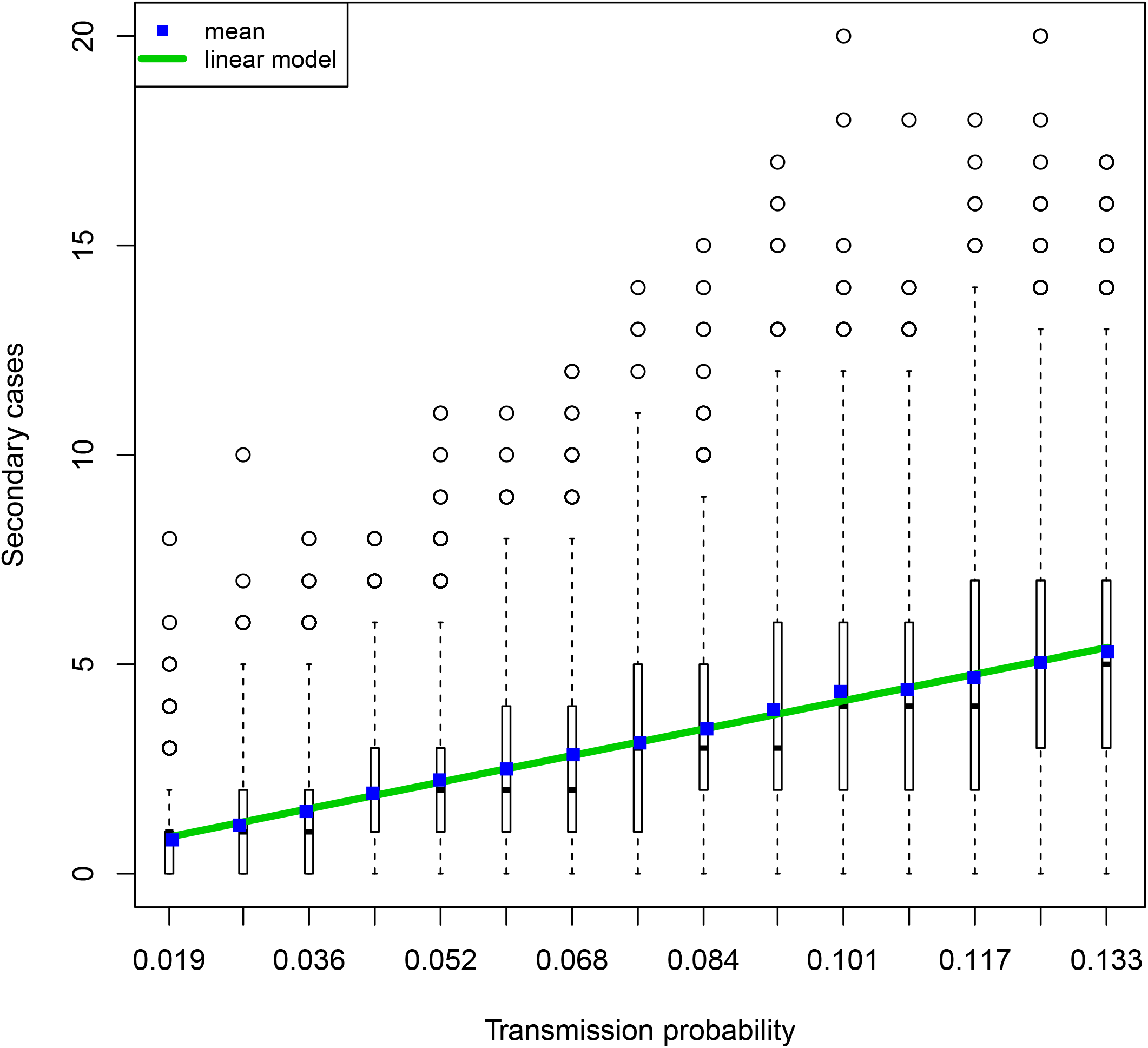
The distribution of secondary cases by transmission probability of 21000 index cases in a susceptible population. The boxplots present the median, quartiles (box), 2.5 and 97.5 percentiles (whiskers) and outliers (circles).

### S6 Hospital admission probability

To estimate time-invariant relative hospital admission probabilities by age, we used the reported hospital admissions up to week 13 (March 30th, 2020) from a Belgian hospital survey [18]. We assumed that hospitals admissions up to week 13 are little affected by the lockdown at the end of week 11 given the mean incubation period of 5.2 days [13], the mean time between symptom onset and hospital admission of 5.6 days [18]. Table S3 present the relative proportion of hospitalized cases by age, averaged for week 11-13. This selection of hospital admission data enables to use a denominator for the number of cases that is not affected by the lockdown, which is the topic of interest in this study. We used the age-specific proportion of symptomatic cases from the simulations to estimate the relationship between the transmission probability and the secondary cases. As such, the calculated relative hospital probabilities are independent from R_0_ and COVID-19 related interventions. We stratified the number of symptomatic cases by 10-year age categories and present them in Table S3. We calculated the relative fraction of symptomatic cases to be hospitalized and a standardized version relative to the oldest age group. These relative proportions are subject to a scaling factor, which we estimated during the parameter estimation procedure as reported in section S7.

**Table S3:** Age-specific proportionality of hospital admission in Belgium based on averaged hospital survey data from week 11-13 and simulated symptomatic cases infected before physical distancing measures occurred. The hospital delay represents the time between symptom onset and hospitalization [18].

| Age category | Reported hospital admissions | Simulated proportion symptomatic cases | Relative fraction | Relative fraction (standardized) | Hospital delay |
| --- | --- | --- | --- | --- | --- |
| 0-9y | 1.4% | 2.0% | 0.72 | 0.091 | 3 days |
| 10-19y | 0.2% | 3.2% | 0.07 | 0.009 | 3 days |
| 20-29y | 1.8% | 5.2% | 0.35 | 0.044 | 7 days |
| 30-39y | 3.9% | 14.7% | 0.26 | 0.033 | 7 days |
| 40-49y | 9.6% | 21.5% | 0.45 | 0.057 | 7 days |
| 50-59 | 15.2% | 25.9% | 0.59 | 0.075 | 7 days |
| 60-69y | 18.5% | 16.4% | 1.13 | 0.143 | 6 days |
| 70-79y | 22.7% | 7.7% | 2.93 | 0.373 | 6 days |
| +80y | 26.7% | 3.4% | 7.87 | 1.000 | 1 day |

### S7 Parameter estimation

We estimated transmission and lockdown characteristics based on reported hospital admissions, initial doubling time and serial seroprevalence data up to May 1st. Afterwards, multiple restrictive measures in Belgium were relaxed, which is the focus of our scenario analysis. From March 14th onward, people who could, were coerced into telework and many businesses, unable to guarantee required hygiene en physical distancing measures had to close. All business-to-consumer (B2C) outlets were interrupted except for ordered home deliveries and shops selling essential goods. All schools were closed except for a low number of children that required day care (e.g., if both parents were not able to telework), so we assumed neither social contacts nor transmission at schools.

We estimated the reduction in social mixing associated with business-to-business (B2B) activities, and in the community (including B2C). An instant reduction from March 14th did not provide a good fit with observed data. Therefore, we included a linear increase in compliance to reduce social contacts. The level of reduction and the duration for this compliance to reach the max is subject of our parameter estimation.

#### Disease and lockdown parameters

- Transmission probability per contact, which we express as R_0_ using the linear model from Section S5.
- The number of introductions into the population and the timing. Since these two parameters are strongly correlated, we choose to fix the introduction date to Monday February 17th, 2020, which is one month prior the study period (lockdown) to allow the model to have some burn-in period without starting within the spring break (February 24-28, 2020).
- Hospital probability scaling factor, to use in combination with the age-specific relative hospital probability values as presented in Table S3.
- Social contact reductions at workplaces and the delay to reach full compliance after March 14th, 2020. We assumed that the compliance increased linearly over time.
- Social contact reductions in the community and the delay to reach full compliance after March 14th, 2020. We assumed that the compliance increased linearly over time.

#### Reference data

- Total hospital admissions per day as reported by the Belgian Health Institute Sciensano [19].
- Doubling time pre-lockdown of 3.1 (2.4-4.4), based on [20]. From each simulation we used the average doubling time between February 24th up to March 8th, 2020.
- Serial seroprevalence of 0.029 (0.023-0.036) on March 19th and 0.060 (0.051-0.071) on April 9th, 2020, based on [21]. These seroprevalence rates are derived from samples collected during one week starting on March 30th and April 20th. We used the midpoint of these sample weeks and assume that the infections took place at least 14 days earlier in order to reflect the minimum time needed to build up IgG antibodies against SARS-CoV-2 that can be detected by ELISA tests [21].

We used the Poisson log-likelihood statistic to assess how well the model describes the reference data. This method is appropriate when dealing with count data, such as the hospital admissions [22]. The log-likelihood when ignoring a constant *log*(*k*_*i*_!) is calculated as

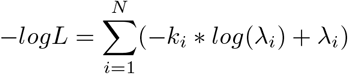

with *k*_*i*_ the observed incidence and *λ*_*i*_ the predicted incidence on time point *i* over the time horizon *N*.

##### Multi-criteria and iterative procedure

Given our interest in hospital admissions, incidence and transmission dynamics, we used three reference outcomes or criteria for which we were able to calculate the log-likelihood. To select optimal parameter combinations, we used the intersection of the 15% best scoring model runs for each criteria. If this intersection contained less than 10 parameter sets, the cutoff was incremented with 2% until at least 10 parameter sets were included in our model ensemble. We started from broad parameter ranges to performed the multi-criteria procedure and used the resulting parameter ensemble to guide the parameter ranges for the subsequent iteration. This process was repeated up to 3 iterations using 1000 Latin Hypercube samples with 5 stochastic realisations. The initial and selected parameter ranges for each iteration are given in Table S4.

The first iteration had almost exclusively impact on the selection of R_0_. During subsequent iterations, the importance of other parameters increased so they could weigh on the ensemble selection. The limited impact of the 3rd iteration made clear we reached a plateau in the parameter estimation procedure. We increased the number of stochastic realisations to 10 to select a final model parameter ensemble. From the latter, which takes all criteria into account, we selected the on average best scoring parameter set according the hospital admission data since this is the model outcome under study for the scenario analyses.

Simulations with more than 1500k cases up to May 1st were stopped and as such excluded from post-processing because this is twice the reported serial seroprevalence in April 2020.

**Figure S12:**
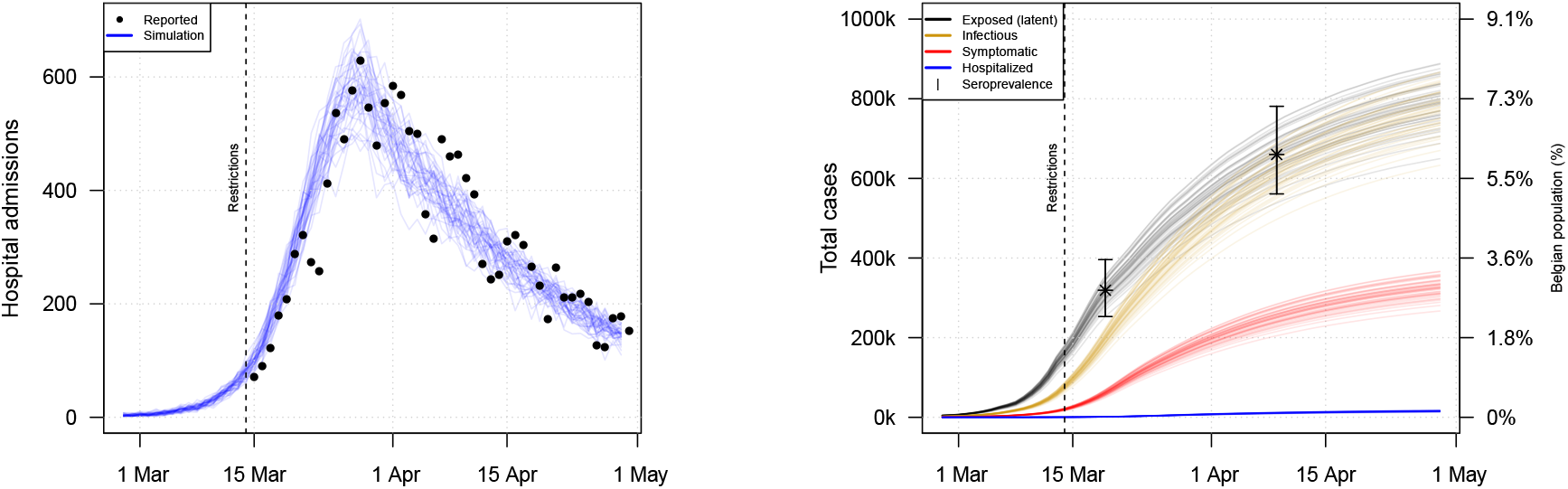
Stochastic simulation results in terms of daily hospital admissions (left) and cumulative incidence of exposed, infectious and symptomatic cases over time (right) using the parameters from the “Final set” in Table S4. The cumulative incidences are presented together with confidence intervals from serial seroprevalence data on March 19th and April 9th based on [21].

**Table S4:** Parameter ranges and observed initial doubling time (min-max) along the estimation procedure and the final model parameter set. The hospital probability factor refers to the +80 age group. For the other age groups, see table S3.

| Parameter | Start | Iteration 1 | Iteration 2 | Iteration 3 | Iteration 4 | Final set |
| --- | --- | --- | --- | --- | --- | --- |
| $R_0^*$ | 1-5 | 3.14 - 3.79 | 3.35 - 3.51 | 3.41 - 3.50 | 3.41 - 3.49 | 3.42 |
| Infected introductions | 200 - 600 | 121 - 556 | 225 - 321 | 233 - 308 | 236 - 307 | 263 |
| Hospital probability factor | 0.05 - 0.9 | 0.20 - 0.81 | 0.26 - 0.31 | 0.35 - 0.47 | 0.35 - 0.46 | 0.4 |
| Contact reduction: workplace | 0.60 - 0.95 | 0.62 - 0.95 | 0.65 - 0.94 | 0.67 - 0.94 | 0.70 - 0.93 | 0.86 |
| Compliance delay: workplace (days) | 5, 6, 7 | 5, 6, 7 | 5, 6, 7 | 5, 6, 7 | 5, 6, 7 | 7 |
| Contact reduction: other | 0.60 - 0.95 | 0.68 - 0.95 | 0.78 - 0.89 | 0.81 - 0.88 | 0.82 - 0.87 | 0.85 |
| Compliance delay: other (days) | 5, 6, 7 | 5, 6, 7 | 5, 6, 7 | 5, 6, 7 | 5, 6, 7 | 7 |
| Latin Hypercube samples | - | 1000 | 1000 | 1000 | 500 | - |
| Stochastic realisations | - | 5 | 5 | 5 | 10 | - |
| Doubling time** (days) | - | 2.81 - 3.43 | 3.04 - 3.21 | 3.01 - 3.16 | 3.00 - 3.18 | 3.09 - 3.17 |

\* $R_0$ is used as input parameter by the reverse calculation to the transmission probability per contact; \*\* model output.

### S8 Age-specific susceptibility

In our default model configuration, the infectiousness of children in the population is limited due to their low probability to be symptomatic and our assumption that asymamptomatic cases are only 50% as infectious [13]. Hence, the probability to be symptomatic for children aged 0-19y is only 7%. To incorporate additional age-specific effects, we re-calibrated our transmission model given that children (0-17y) are only halve as susceptible compared to adults (+18y). This change in disease characteristics required an update of the relationship between the transmission probability per contact and the number of secondary cases (i.e. the reproduction number). The procedure as described in Section S5 has been applied with the altered disease characteristics and resulted into:

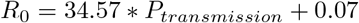

and the inverse:

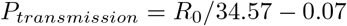

We also re-estimated the relative hospital probability by age since the proportion of symptomatic cases before the lockdown changed. The results are presented in Table S5. Finally, we repeated the iterative parameter estimation procedure as described in Section S7, and present the results in Table S6. The resulting model fit is presented in Figure S13.

Simulations for the baseline scenario with age-specific susceptibility in terms of hospital admissions and reproduction number (Figure S14) are similar to the results presented in the main text (Figure 1). Also the simulations with household bubbles, contact tracing and a combined strategy scenario analyses are not much affected by the susceptibility assumption (Figure S15). For strategies involving school re-opening, the effect is substantial as discussed in the main text.

**Table S5:** Age-specific proportionally of hospital admission when children (0-17y) are only 50% as susceptible compared to adults (+18y). The hospital delay represents the time between symptom onset and hospitalization [18].

| Age category | Reported hospital admissions | Simulated symptomatic cases | Fraction | Fraction (standardized) | Hospital delay |
| --- | --- | --- | --- | --- | --- |
| 0-9y | 1.4% | 1% | 1.71 | 0.127 | 3 days |
| 10-19y | 0.2% | 2% | 0.12 | 0.009 | 3 days |
| 20-29y | 1.8% | 6% | 0.33 | 0.024 | 7 days |
| 30-39y | 3.9% | 15% | 0.26 | 0.019 | 7 days |
| 40-49y | 9.6% | 22% | 0.44 | 0.033 | 7 days |
| 50-59 | 15.2% | 27% | 0.56 | 0.041 | 7 days |
| 60-69y | 18.5% | 17% | 1.10 | 0.081 | 6 days |
| 70-79y | 17.0% | 7.7% | 2.20 | 0.163 | 6 days |
| +80y | 45.8% | 3.4% | 13.51 | 1.000 | 1 day |

**Table S6:** Parameter ranges and observed initial doubling time (min-max) along the estimation procedure with age-specific susceptibility and final model parameter set. The hospital probability factor refers to the +80 age group. For the other age groups, see table S5.

| Parameter | Start | Iteration 1 | Iteration 2 | Iteration 3 | Iteration 4 | Final set |
| --- | --- | --- | --- | --- | --- | --- |
| $R_0^*$ | 1-5 | 3.00 - 3.48 | 3.35 - 3.45 | 3.36 - 3.43 | 3.37 - 3.45 | 3.37 |
| Infected introductions | 200-600 | 206 - 593 | 210 - 322 | 230 - 389 | 217 - 279 | 225 |
| Hospital probability factor | 0.05 - 0.9 | 0.14 - 0.75 | 0.24 - 0.44 | 0.36 - 0.47 | 0.31 - 0.38 | 0.35 |
| Contact reduction: workplace | 0.60 - 0.95 | 0.61 - 0.94 | 0.64 - 0.90 | 0.69 - 0.92 | 0.68 - 0.89 | 0.76 |
| Compliance delay: workplace (days) | 5, 6, 7 | 5, 6, 7 | 5, 6, 7 | 5, 6, 7 | 5, 6, 7 | 6 |
| Contact reduction: other | 0.60 - 0.95 | 0.70 - 0.94 | 0.83 - 0.89 | 0.83 - 0.92 | 0.83 - 0.88 | 0.86 |
| Compliance delay: other (days) | 5, 6, 7 | 5, 6, 7 | 5, 6, 7 | 5, 6, 7 | 5, 6, 7 | 7 |
| Latin Hypercube samples | - | 1000 | 1000 | 1000 | 500 |  |
| Stochastic realisations per set | - | 5 | 5 | 5 | 10 |  |
| Doubling time** (days) | - | 3.03 - 3.54 | 3.04 - 3.19 | 3.09 - 3.11 | 3.00 - 3.20 | 3.09 - 3.19 |

\* $R_0$ is used as input parameter by the reverse calculation to a transmission probability per contact; \*\* model output.

**Figure S13:**
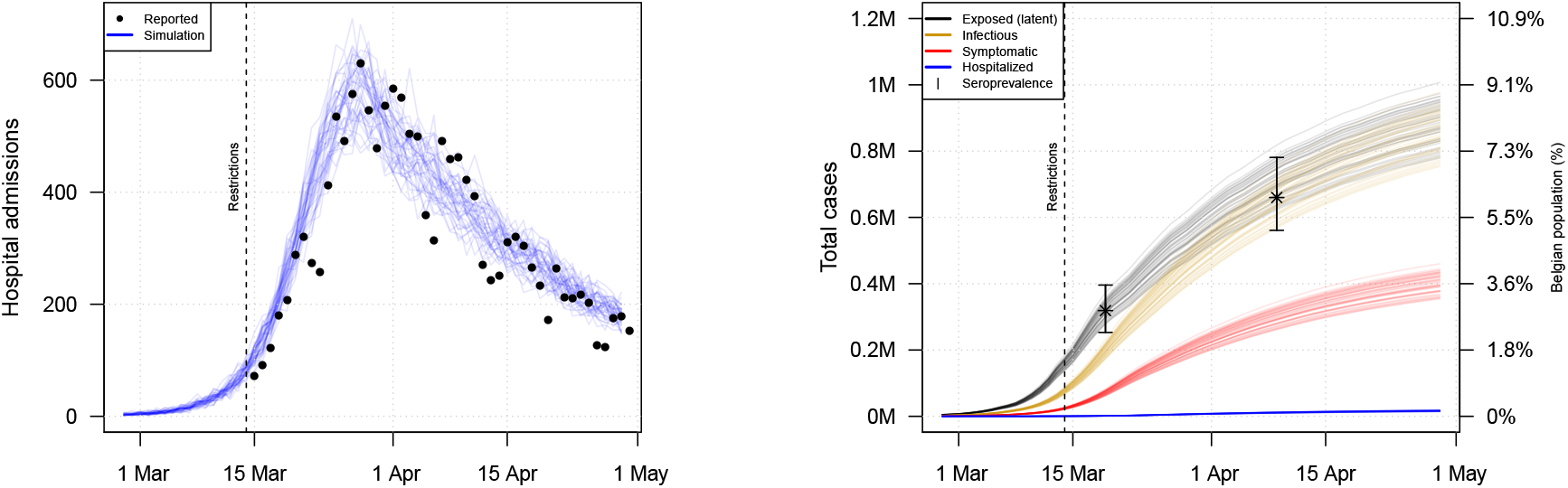
Stochastic simulation results with age-specific susceptibility in terms of daily hospital admissions (left) and cumulative incidence of exposed, infectious and symptomatic cases over time (right) based on the parameters from the “Final set” in Table S6. The cumulative incidences are presented together with confidence intervals from serial seroprevalence data on March 19th and April 9th based on [21].

**Figure S14:**
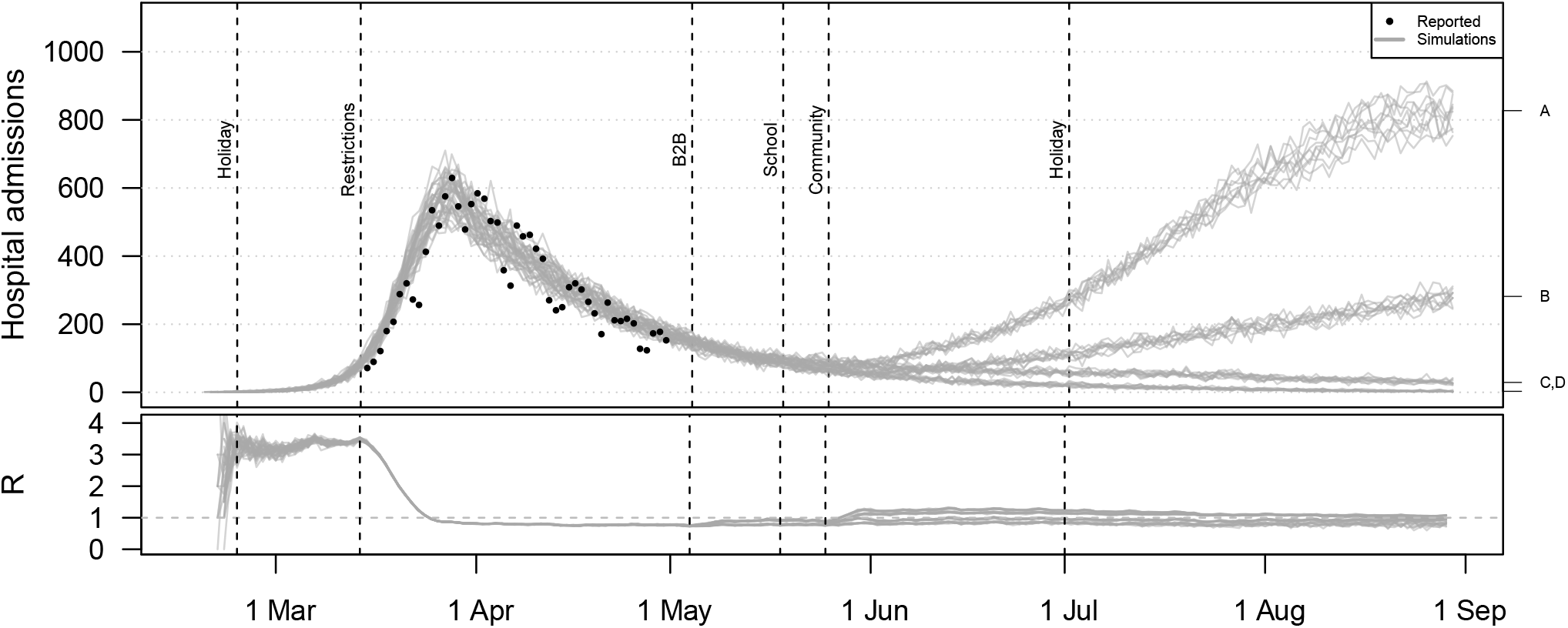
Hospital admissions and effective reproduction number (R) from the baseline scenario including 4 mixing assumptions with age-specific susceptibility. All simulations include social restrictions from March 14th and the partial school reopening in May. For the B2B, the social mixing after the lockdown is assumed to double from the indicated point in time (indicated with A and C) or to remain constant (B,D). Social mixing in the community is assumed to double (A,B) or to remain constant (C,D).

**Figure S15:**
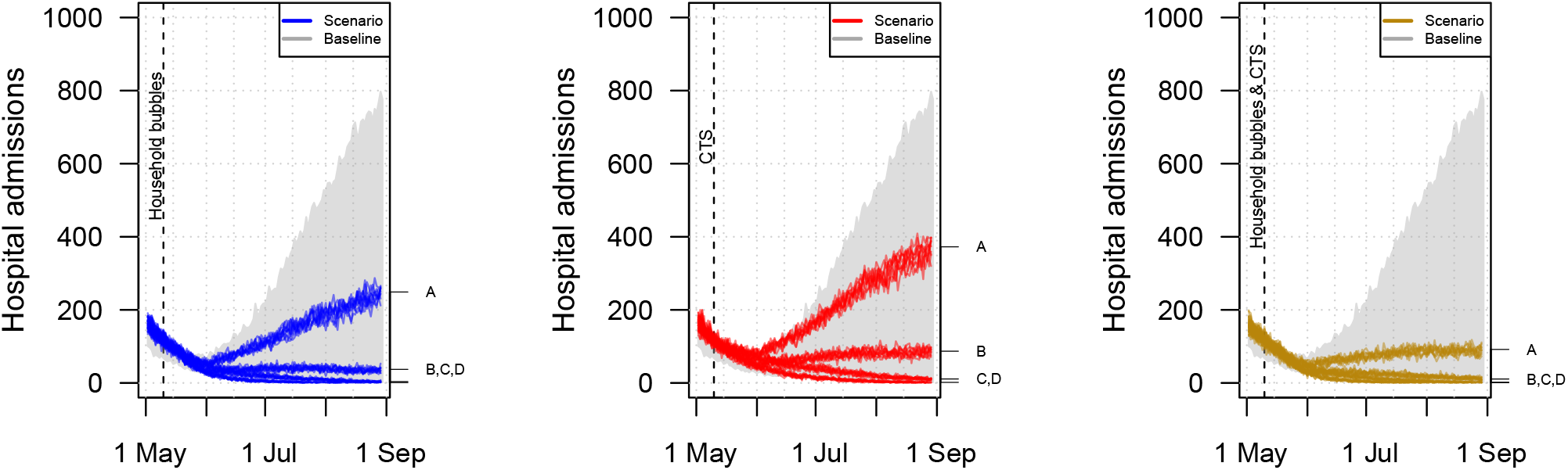
Hospital admissions over time when community mixing occurs in household bubbles (left), a contact tracing strategy (CTS) is in place (center), or both (right) with age-specific susceptibility. All scenarios are based on the same natural disease history and quantitative mixing assumptions but differ from the baseline in terms of the network structure and application of contact tracing from the given point in time. The mixing assumptions A,B,C,D are explained in the caption of Figure S14.

### S9 Scenario definitions

We defined deconfinement scenarios by combinations of location-specific social mixing relative to pre-pandemic observations. By varying percentages of contacts at different locations, we implicitly assumed people either make fewer contacts compared to the pre-pandemic situation or the contacts they made were less likely to lead to transmission. For example, if we increased social mixing at workplaces from 25% to 50%, we predict the impact of “what if the risk to acquire infection at work doubled compared to during lockdown, though still 50% less than in pre-pandemic times”. To account for structural uncertainty with respect to B2B-related social mixing, we included 25% and 50% of the social contacts rates prior lockdown. For community-related mixing, we used 15% and 30% of the social contact rates prior lockdown. By combining these B2B and community mixing patterns, we ended up with four social mixing parameter sets and we ran 10 stochastic realisations per parameter set. Table S7 contains temporal aspects of the scenarios and the age-specific “school*” program based on the Belgian regulations.

**Table S7:** Overview of lockdown measures and gradual relief in the scenario analyses.

| What? | Timing? |
| --- | --- |
| Start restriction measures by closing schools, universities, cultural activities, bars and restaurants. Four days later, additional measures were taken to allow only work-related transport of essential workers, and teleworking made the norm. | March 14th |
| Re-start business-to-business (B2B) | May 4th |
| Social mixing in household bubbles | May 11th |
| Start contact tracing strategies (CTS) | May 11th |
| Re-open school* for 0-2year (daycare) & 6-7year (1st and 2nd grade primary school) | May 18th, 4 days / week |
| Re-open school* for 11year olds (6th grade primary school) | May 18th, 2 days / week |
| Re-open school* for 17year olds (6th grade secondary school) | May 18th, 1 day / week |
| Re-open schools for 0-5year, 0-11year, 0-17year | May 18th, 5 days / week |
| Increasing leisure, B2C and other social contacts (community) | May 25th |

\* Age-specific school-reopening as stated by the Belgian government on April 24th.

### S10 Robustness analyses

There is no golden standard for the number of stochastic realisations for this type of stochastic simulator. The results in the main text are based on 10 realisations, but we present here a robustness analysis for the main scenarios (baseline, household bubbles, CTS and combined strategy) based on 20, 40 and 80 realisations. We did observe stochastic changes for the projected hospital admissions over time (Figure S16 and S17) but no differences on the averages and average differences in terms of total hospital admissions (Figure S18) with an increasing number of stochastic realisations.

**Figure S16:**
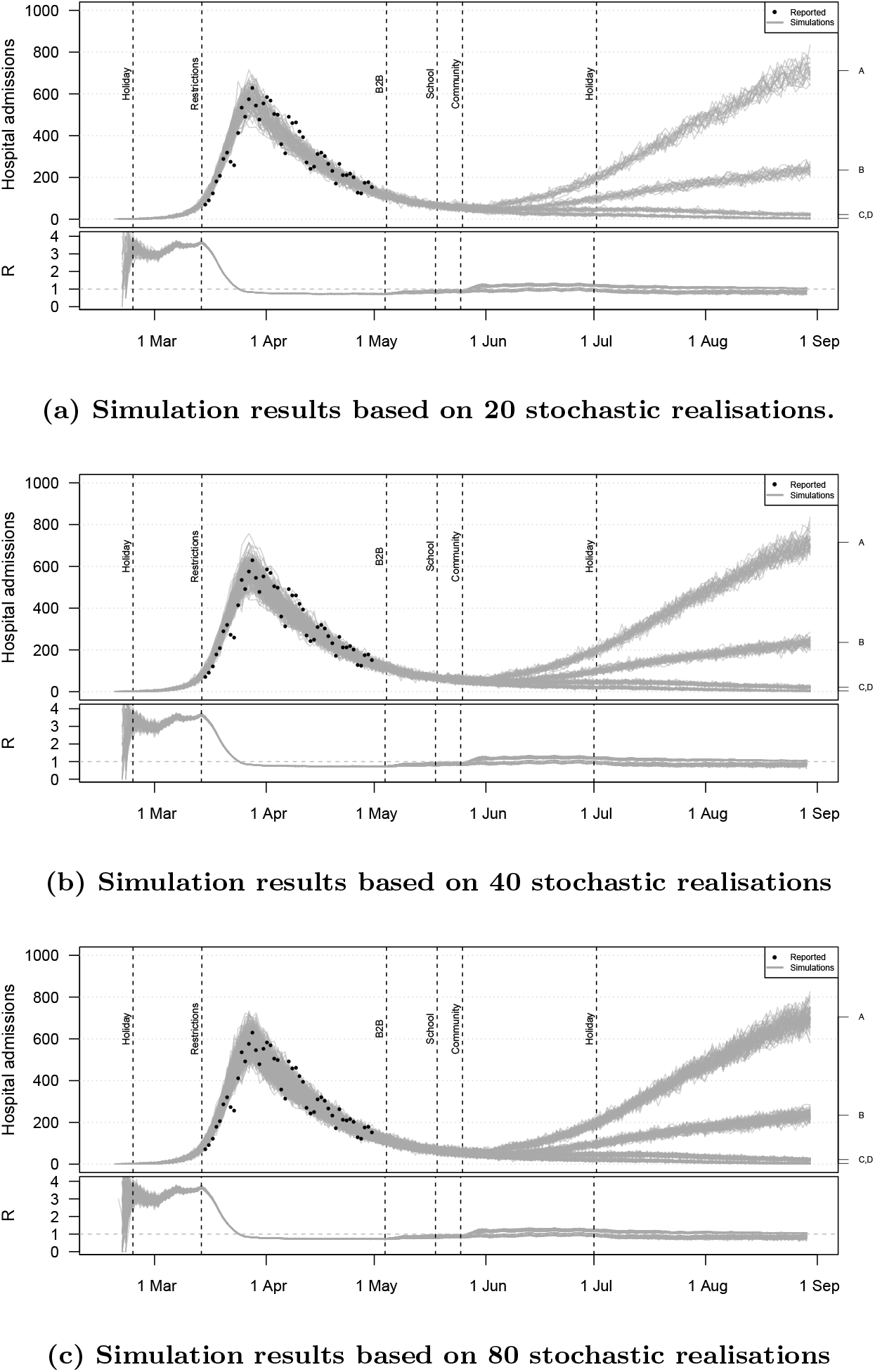
Hospital admissions over time for the baseline scenario based on 20, 40 and 80 stochastic realisations per social contact assumption. All simulations include social restrictions from March 14th and the partial school reopening in May. For the B2B, the social mixing after the lockdown is assumed to double from the indicated point in time (indicated with A and C) or to remain constant (B,D). Social mixing in the community is assumed to double (A,B) or to remain constant (C,D). The dots present the reported hospital admissions for Belgium.

**Figure S17:**
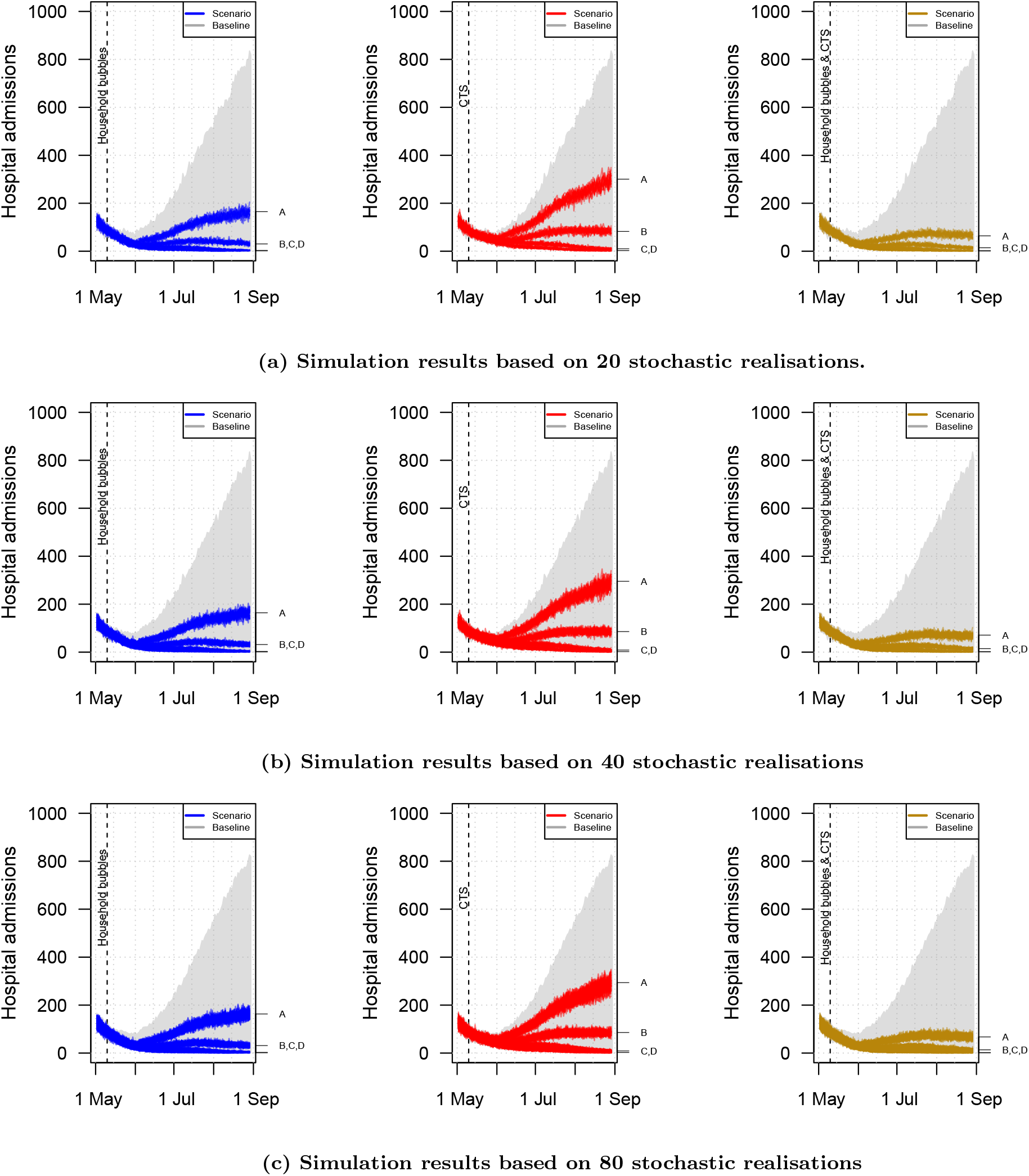
Hospital admissions over time when community mixing occurs in household bubbles (left), a contact tracing strategy (CTS) is in place (center), or both (right) based on 20, 40 and 80 stochastic realisations per social contact assumption. All scenarios are based on the same natural disease history and quantitative mixing assumptions but differ from the baseline in terms of the network structure and application of contact tracing from the given point in time. The mixing assumptions A,B,C,D are explained in the caption of Figure S16..

**Figure S18:**
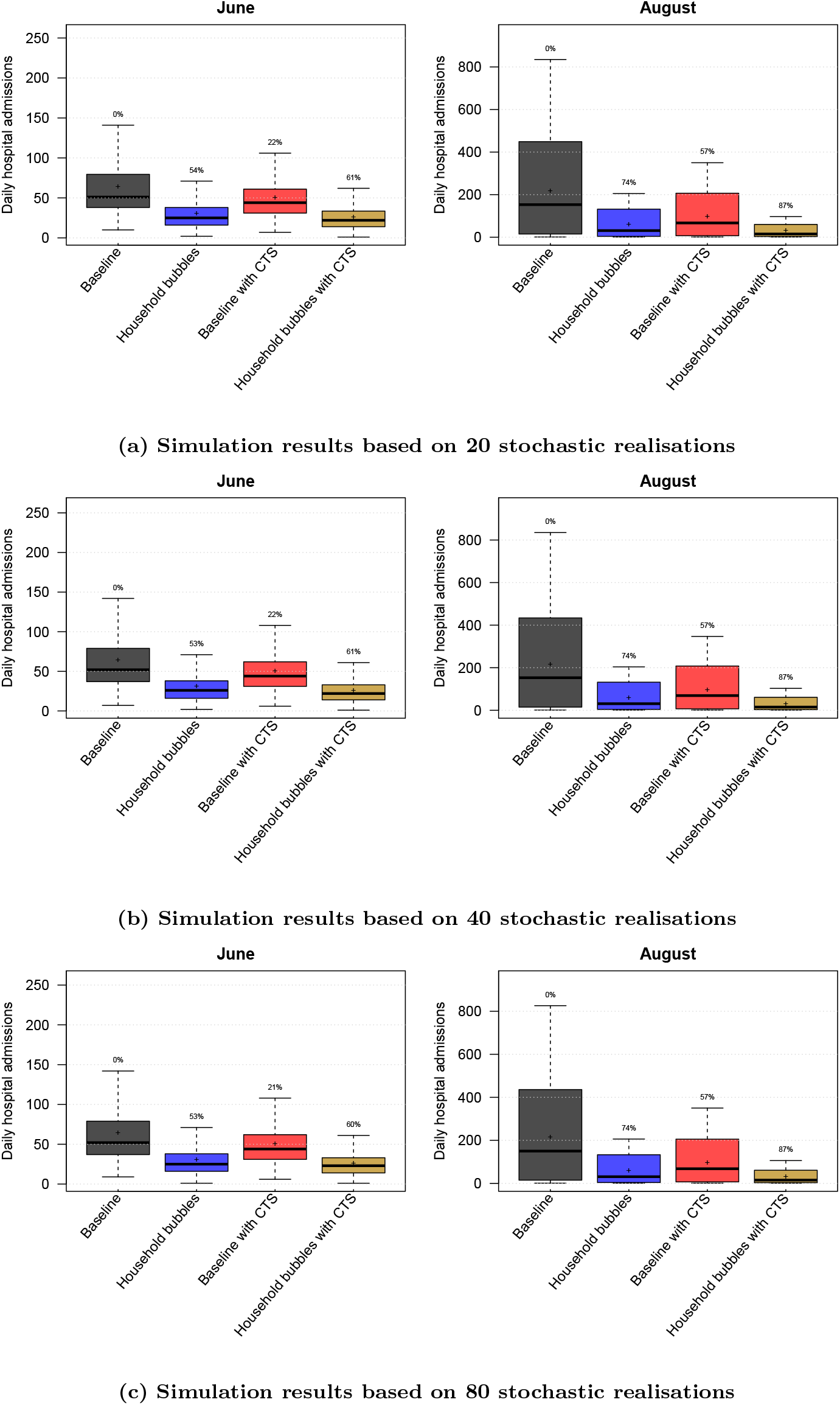
Distribution of the daily hospital admissions by June and August per scenario based on 20, 40 and 80 stochastic realisations per social contact assumption. The results are presented as the median (line), quartiles (box), 2.5 and 97.5 percentiles (whiskers) and average (cross) of the scenario results including social mixing uncertainty and stochastic effects. The percentage on top of the whiskers indicates relative reduction of the scenario average with respect to the baseline. CTS: contact tracing strategy.

### S11 Ensemble analyses

The scenario analyses presented in the main text are based on single parameter estimations from the iterative parameter estimation procedure (Section S7). Given the correlated nature of different model parameters, different combinations can give a similar fit for the first wave, but lead to different outcomes in terms of the scenario analyses. To endorse our results, we ran the main scenarios with all parameter sets from the model ensemble from the 4th iteration with and without age-specific susceptibility. The resulting hospital admissions over time in Figure S19 and S20 include more variation the hospital admissions over time. The average reductions in hospital admissions up to August, as presented in Figure S21, are similar to the results presented in our main analysis (Figure 2). We conclude that our results in terms of aggregated statistics based on our most optimal parameter set remain valid when we include parameter uncertainty.

**Figure S19:**
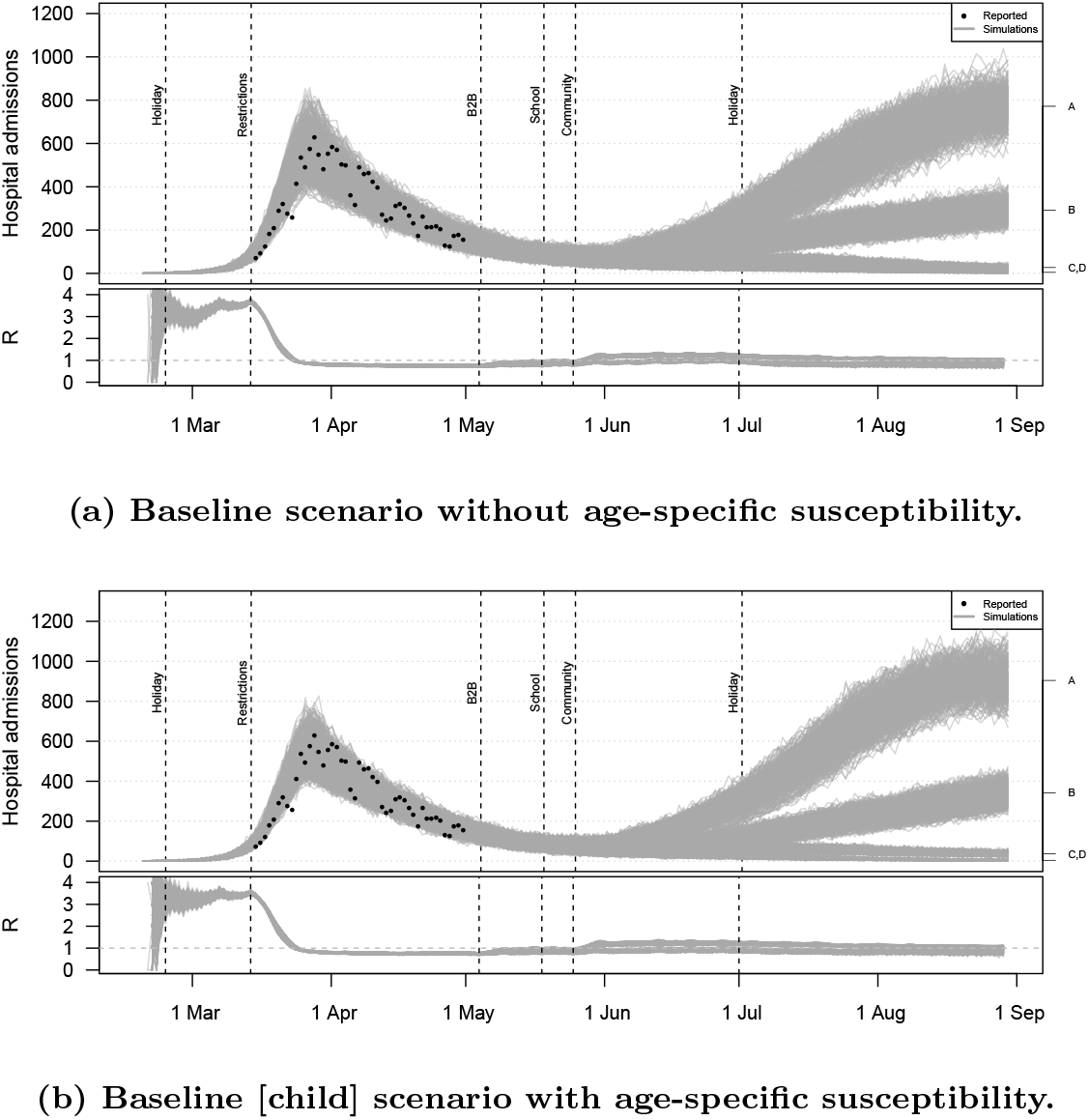
Hospital admissions over time for the baseline scenario based on the model parameter ensemble. All simulations include social restrictions from March 14th and the partial school reopening in May. For the B2B, the social mixing after the lockdown is assumed to double from the indicated point in time (indicated with A and C) or to remain constant (B,D). Social mixing in the community is assumed to double (A,B) or to remain constant (C,D). The dots present the reported hospital admissions for Belgium.

**Figure S20:**
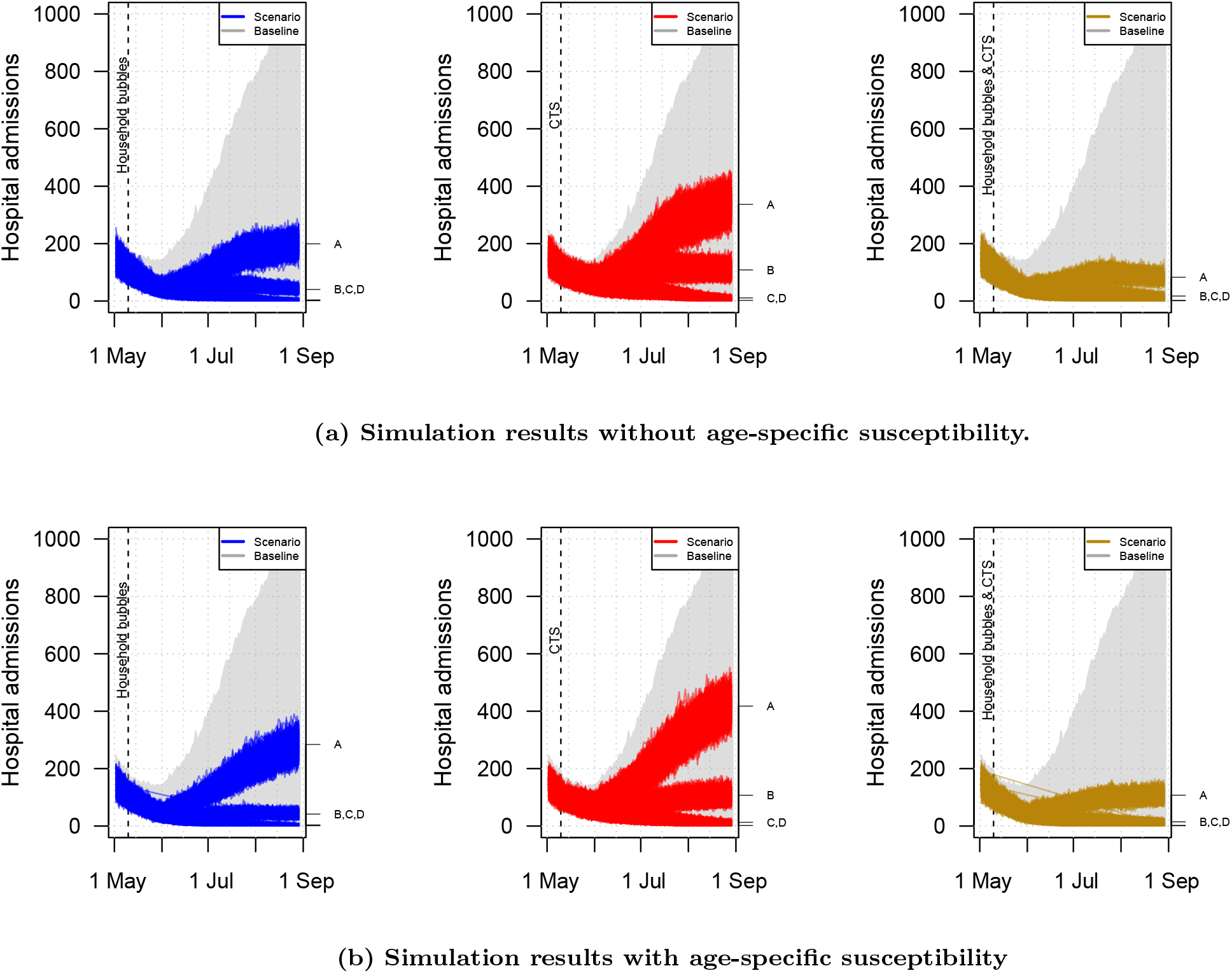
Hospital admissions over time when community mixing occurs in household bubbles (left), a contact tracing strategy (CTS) is in place (center), or both (right) based on the model parameter ensemble. All scenarios are based on the same natural disease history and quantitative mixing assumptions but differ from the baseline in terms of the network structure and application of contact tracing from the given point in time. The mixing assumptions A,B,C,D are explained in the caption of Figure S19.

**Figure S21:**
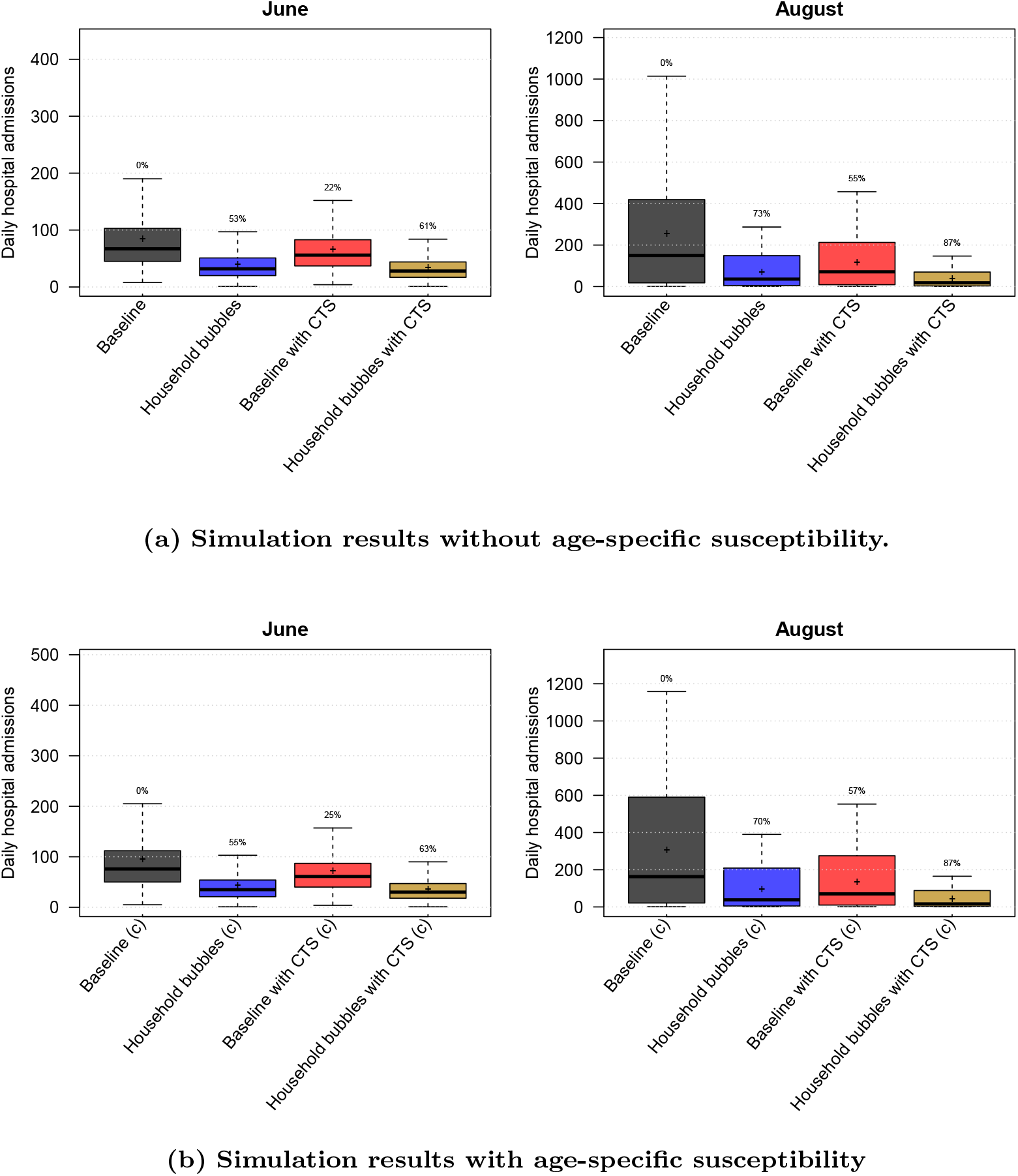
Distribution of the daily hospital admissions by June and August per scenario based on the model parameter ensemble. The results are presented as the median (line), quartiles (box), 2.5 and 97.5 percentiles (whiskers) and average (cross) of the scenario results including social mixing uncertainty and stochastic effects. The percentage on top of the whiskers indicates relative reduction of the scenario average with respect to the baseline. CTS: contact tracing strategy.

### S12 Platform and technical details

STRIDE is open source software (https://github.com/lwillem/stride_covid19_v1) and implemented in C++. The software can be compiled on Linux and Mac OSX platforms with a recent version of a C++ compiler. To build and install STRIDE, the following tools need to be available on the system: GNU g++ or LLVM clang++ compiler, make, CMake and Boost. To generate documentation, Doxygen and LaTeX are required. The build system for STRIDE uses the CMake tool to compile and install the software almost platform independent. More info is provided in the user manual on Github.

Our model is optimized with a switch to evaluate only the social contacts of the infectious individuals instead of matching every individual with all others within one location. More details on our model optimisations are provided in [1].

The modelling project contains regression tests in the Google Test suite embedded in a Travis continuous integration environment. The test environment has been created and maintained during model development for influenza and measles, and additional unit tests were added during this analysis. We implemented a baseline COVID-19 test and also benchmark physical distancing, household bubbles and contact tracing scenarios.

We implemented an “rSTRIDE” framework in R to handle the design of experiments, to run all parameter sets and analyse the output. Different serial STRIDE simulations are run in parallel using the “doParallel” package and the aggregation of summary statistics, prevalence, incidence and social contacts is automated. The synthetic population of 11 million individuals is computed once using R and loaded onto the C++ simulator for every new simulation.

All results presented in this manuscript are generated on the VSC-cluster “Vaughan”, a NEC system consisting of 104 nodes with two 32-core AMD Epyc 7452 Rome generation CPUs connected through a HDR100 InfiniBand network. All nodes have 256 GB RAM. One single run from the baseline scenario (196 days) required *±*20 minutes.

## References

[1] Keeling, M.J., Hollingsworth, T.D., Read, J.M.: The efficacy of contact tracing for the containment of the 2019 novel coronavirus (COVID-19). J Epidemiol Communinty Health 74, 861–866 (2020)

[2] Willem, L., Verelst, F., Bilcke, J., Hens, N., Beutels, P.: Lessons from a decade of individual-based models for infectious disease transmission: a systematic review (2006-2015). BMC infect dis 17(1), 612 (2017)

[3] Kerr, C.C., Stuart, R.M., Mistry, D., Abeysuriya, R.G., Hart, G., Rosenfeld, K., Selvaraj, P., Nunez, R.C., Hagedorn, B., George, L., et al.: Covasim: an agent-based model of COVID-19 dynamics and interventions. medRxiv (2020)

[4] Verelst, F., Willem, L., Beutels, P.: Behavioural change models for infectious disease transmission: a systematic review (2010-2015). J R Soc Interface 13(125), 20160820 (2016)

[5] Hoang, T.V., Coletti, P., Melegaro, A., Wallinga, J., Grijalva, C.G., Edmunds, J.W., Beutels, P., Hens, N.: A systematic review of social contact surveys to inform transmission models of close-contact infections. Epidemiology 30(5), 723–736 (2019)

[6] Willem, L., Hoang, V.T., Funk, S., Coletti, P., Beutels, P., Hens, N.: SOCRATES: an online tool leveraging a social contact data sharing initiative to assess mitigation strategies for COVID-19. BMC Res Notes 13(1), 293 (2020)

[7] Wallinga, J., Teunis, P., Kretzschmar, M.: Using data on social contacts to estimate age-specific transmission parameters for respiratory-spread infectious agents. Am J Epidemiol 164(10), 936–944 (2006)

[8] Goeyvaerts, N., Hens, N., Ogunjimi, B., Aerts, M., Shkedy, Z., Van Damme, P., Beutels, P.: Estimating infectious disease parameters from data on social contacts and serological status. J R Stat Soc Ser C Appl Stat 59(2), 255–277 (2010)

[9] Sciensano, Belgium: COVID-19 - Epidemiologische situatie. https://epistat.wiv-isp.be/covid/ (2020)

[10] Willem, L., Stijven, S., Tijskens, E., Beutels, P., Hens, N., Broeckhove, J.: Optimizing agent-based transmission models for infectious diseases. BMC Bioinformatics 16, 183 (2015)

[11] Kuylen, E., Stijven, S., Broeckhove, J., Willem, L.: Social contact patterns in an individual-based simulator for the transmission of infectious diseases (Stride). In: ICCS, pp. 2438–2442 (2017)

[12] Kuylen, E., Willem, L., Broeckhove, J., Beutels, P., Hens, N.: Clustering of susceptible individuals within households can drive measles outbreaks: an individual-based model exploration. Sci Rep 10(19645) (2020)

[13] Willem, L., Van Kerckhove, K., Chao, D.L., Hens, N., Beutels, P.: A nice day for an infection? Weather conditions and social contact patterns relevant to influenza transmission. PLoS One 7(11) (2012)

[14] Kifle, Y.W., Goeyvaerts, N., Van Kerckhove, K., Willem, L., Kucharski, A., Faes, C., Leirs, H., Hens, N., Beutels, P.: Animal ownership and touching enrich the context of social contacts relevant to the spread of human infectious diseases. PloS One 10(7), 0133461 (2015)

[15] Van Hoang, T., Coletti, P., Kiffle, Y.W., Van Kerckhove, K., Vercruysse, S., Willem, L., Beutels, P., Hens, N.: Close contact infection dynamics over time: insights from a second large-scale social contact survey in Flanders, Belgium, in 2010-2011. medRxiv (2020)

[16] Van Kerckhove, K., Hens, N., Edmunds, W.J., Eames, K.T.: The impact of illness on social networks: implications for transmission and control of influenza. Am J epidemiol 178(11), 1655–1662 (2013)

[17] Pellis, L., Scarabel, F., Stage, H.B., Overton, C.E., Chappell, L.H., Lythgoe, K.A., Fearon, E., Bennett, E., Curran-Sebastian, J., Das, R., et al.: Challenges in control of Covid-19: short doubling time and long delay to effect of interventions. 2004.00117 (2020)

[18] Herzog, S., De Bie, J., Abrams, S., Wouters, I., Ekinci, E., Patteet, L., Coppens, A., De Spiegeleer, S., Beutels, P., Van Damme, P., Hens, N., Theeten, H.: Seroprevalence of IgG antibodies against SARS coronavirus 2 in Belgium: a serial prospective cross-sectional nationwide study of residual samples. medRxiv (2020)

[19] Liu, Y., Gayle, A.A., Wilder-Smith, A., Rocklöv, J.: The reproductive number of COVID-19 is higher compared to SARS coronavirus. J Travel Med 1, 4 (2020)

[20] Abrams, S., Wambua, J., Santermans, E., Willem, L., Kuylen, E., Coletti, P., Libin, P., Faes, C., Petrof, O., Herzog, S.A., SIMID COVID19 team, Beutels, P., Hens, N.: Modeling the early phase of the Belgian COVID-19 epidemic using a stochastic compartmental model and studying its implied future trajectories. medRxiv (2020)

[21] Coletti, P., Libin, P., Petrof, O., Willem, L., Abrams, S., Faes, C., Wambua, J., Kuylen, E., Beutels, P., Hens, N.: A data-driven metapopulation model for the Belgian COVID-19 epidemic: assessing the impact of lockdown and exit strategies. medRxiv (2020)

[22] Prather, K.A., Wang, C.C., Schooley, R.T.: Reducing transmission of SARS-CoV-2. Science 368(6498), 1422–1424 (2020)

[23] Davies, N.G., Klepac, P., Liu, Y., Prem, K., Jit, M., Eggo, R.M., CMMID-COVID-19 Working Group, et al.: Age-dependent effects in the transmission and control of COVID-19 epidemics. Nat Med (2020)

[24] Leng, T., White, C., Hilton, J., Kucharski, A., Pellis, L., Stage, H., Davies, N., CMMID-COVID-19 Working Group, M.J. Keeling, Flasche, S.: The effectiveness of social bubbles as part of a Covid-19 lockdown exit strategy, a modelling study. Technical report (2020)

[25] Nishiura, H., Linton, N.M., Akhmetzhanov, A.R.: Serial interval of novel coronavirus (COVID-19) infections. Int J Infect Dis 93, 284–286 (2020)

[26] Kucharski, A.J., Klepac, P., Conlan, A., Kissler, S.M., Tang, M., Fry, H., Gog, J., Edmunds, J., CMMID-COVID-19 Working Group, et al.: Effectiveness of isolation, testing, contact tracing and physical distancing on reducing transmission of SARS-CoV-2 in different settings. Lancet Infect Dis (2020)

[27] Kretzschmar, M.E., Rozhnova, G., Bootsma, M.C., van Boven, M., van de Wijgert, J.H., Bonten, M.J.: Impact of delays on effectiveness of contact tracing strategies for covid-19: a modelling study. Lancet Public Health 5(8), 452–459 (2020)

[28] Libin, P.J.K., Willem, L., Verstraeten, T., Torneri, A., Vanderlocht, J., Hens, N.: Assessing the feasibility and effectiveness of household-pooled universal testing to control COVID-19 epidemics. medRxiv (2020)

[29] Cauchemez, S., Valleron, A.-J., Boelle, P.-Y., Flahault, A., Ferguson, N.M.: Estimating the impact of school closure on influenza transmission from Sentinel data. Nature 452(7188), 750–754 (2008)

[30] Hens, N., Ayele, G.M., Goeyvaerts, N., Aerts, M., Mossong, J., Edmunds, J.W., Beutels, P.: Estimating the impact of school closure on social mixing behaviour and the transmission of close contact infections in eight European countries. BMC Infect Dis 9(1), 187 (2009)

[31] Chao, D.L., Oron, A.P., Srikrishna, D., Famulare, M.: Modeling layered non-pharmaceutical interventions against SARS-CoV-2 in the United States with Corvid. medRxiv (2020)

[32] Eubank, S., Eckstrand, I., Lewis, B., Venkatramanan, S., Marathe, M., Barrett, C.: Commentary on Ferguson, et al.,”Impact of non-pharmaceutical interventions (NPIs) to reduce COVID-19 mortality and healthcare demand”. Bull Math Biol 82, 1–7 (2020)

[33] Panovska-Griffiths, J., Kerr, C.C., Stuart, R.M., Mistry, D., Klein, D.J., Viner, R.M., Bonell, C.: Determining the optimal strategy for reopening schools, the impact of test and trace interventions, and the risk of occurrence of a second COVID-19 epidemic wave in the UK: a modelling study. Lancet Child Adolesc Health 4(11), 817–827 (2020)

[34] Torneri, A., Libin, P., Vanderlocht, J., Vandamme, A.-M., Neyts, J., Hens, N.: A prospect on the use of antiviral drugs to control local outbreaks of COVID-19. BMC Medicine 18(1), 191 (2020)

[35] Franco, N.: Covid-19 belgium: Extended seir-qd model with nursery homes and long-term scenarios-based forecasts from school opening. arXiv preprint 2009.03450 (2020)

## References (Supplementary Material)

[1] Willem, L., Stijven, S., Tijskens, E., Beutels, P., Hens, N., Broeckhove, J.: Optimizing agent-based transmission models for infectious diseases. BMC Bioinformatics 16, 183 (2015)

[2] Kuylen, E., Stijven, S., Broeckhove, J., Willem, L.: Social contact patterns in an individual-based simulator for the transmission of infectious diseases (Stride). In: ICCS, pp. 2438–2442 (2017)

[3] Kuylen, E., Willem, L., Broeckhove, J., Beutels, P., Hens, N.: Clustering of susceptible individuals within households can drive measles outbreaks: an individual-based model exploration. Sci Rep 10(19645) (2020)

[4] EUROSTAT: Your Key to European Statistics. https://ec.europa.eu/eurostat/data/

[5] Kind en Gezin. Vlaamse Overheid: Opvangadressen [status 2019-09-24]. https://www.kindengezin.be/toepassingen/zoekopvang.jsp (2019)

[6] Onderwijs Vlaanderen: Publicaties omkadering lestijden gewoon basisonderwijs 2012-2013. https://www.agodi.be/nieuwe-omkadering-basisonderwijs (2013)

[7] Willem, L., Van Kerckhove, K., Chao, D.L., Hens, N., Beutels, P.: A nice day for an infection? Weather conditions and social contact patterns relevant to influenza transmission. PLoS One 7(11) (2012)

[8] Kifle, Y.W., Goeyvaerts, N., Van Kerckhove, K., Willem, L., Kucharski, A., Faes, C., Leirs, H., Hens, N., Beutels, P.: Animal ownership and touching enrich the context of social contacts relevant to the spread of human infectious diseases. PloS One 10(7), 0133461 (2015)

[9] Van Hoang, T., Coletti, P., Kiffle, Y.W., Van Kerckhove, K., Vercruysse, S., Willem, L., Beutels, P., Hens, N.: Close contact infection dynamics over time: insights from a second large-scale social contact survey in Flanders, Belgium, in 2010-2011. medRxiv (2020)

[10] Goeyvaerts, N., Santermans, E., Potter, G., Torneri, A., Van Kerckhove, K., Willem, L., Aerts, M., Beutels, P., Hens, N.: Household members do not contact each other at random: implications for infectious disease modelling. Proc R Soc B 285(1893), 20182201 (2018)

[11] Funk, S.: socialmixr: Social Mixing Matrices for Infectious Disease Modelling. The Comprehensive R Archive Network (2020)

[12] Willem, L., Hoang, V.T., Funk, S., Coletti, P., Beutels, P., Hens, N.: SOCRATES: an online tool leveraging a social contact data sharing initiative to assess mitigation strategies for COVID-19. BMC Res Notes 13(1), 293 (2020)

[13] Li, Q., Guan, X., Wu, P., Wang, X., Zhou, L., Tong, Y., Ren, R., Leung, K.S.M., Lau, E.H.Y., et al.: Early transmission dynamics in Wuhan, China, of novel coronavirus–infected pneumonia. N Engl J Med 382(13), 1199–1207 (2020)

[14] Wu, J.T., Leung, K., Bushman, M., Kishore, N., Niehus, R., de Salazar, P.M., Cowling, B.J., Lipsitch, M., Leung, G.M.: Estimating clinical severity of COVID-19 from the transmission dynamics in Wuhan, China. Nat Med, 1–5 (2020)

[15] He, X., Lau, E.H., Wu, P., Deng, X., Wang, J., Hao, X., Lau, Y.C., Wong, J.Y., Guan, Y., Tan, X., et al.: Temporal dynamics in viral shedding and transmissibility of COVID-19. Nat Med 26(5), 672–675 (2020)

[16] Lourenco, J., Paton, R., Ghafari, M., Kraemer, M., Thompson, C., Simmonds, P., Klenerman, P., Gupta, S.: Fundamental principles of epidemic spread highlight the immediate need for large-scale serological surveys to assess the stage of the SARS-CoV-2 epidemic. MedRxiv (2020)

[17] Li, R., Pei, S., Chen, B., Song, Y., Zhang, T., Yang, W., Shaman, J.: Substantial undocumented infection facilitates the rapid dissemination of novel coronavirus (SARS-CoV-2). Science 368(6490), 489–493 (2020)

[18] Faes, C., Abrams, S., Van Beckhoven, D., Meyfroidt, G., Vlieghe, E., Hens, N.: Time between symptom onset, hospitalisation and recovery or death: Statistical analysis of Belgian COVID-19 patients. Int J Environ Res Public Health 17(20), 7560 (2020)

[19] Sciensano, Belgium: COVID-19 - Epidemiologische situatie. https://epistat.wiv-isp.be/covid/ (2020)

[20] Pellis, L., Scarabel, F., Stage, H.B., Overton, C.E., Chappell, L.H., Lythgoe, K.A., Fearon, E., Bennett, E., Curran-Sebastian, J., Das, R., et al.: Challenges in control of Covid-19: short doubling time and long delay to effect of interventions. 2004.00117 (2020)

[21] Herzog, S., De Bie, J., Abrams, S., Wouters, I., Ekinci, E., Patteet, L., Coppens, A., De Spiegeleer, S., Beutels, P., Van Damme, P., Hens, N., Theeten, H.: Seroprevalence of IgG antibodies against SARS coronavirus 2 in Belgium: a serial prospective cross-sectional nationwide study of residual samples. medRxiv (2020)

[22] Hilbe, J.: Modeling Count Data., (2014)

